# Independent Inheritance of Cognition and Bipolar Disorder in a Family Sample

**DOI:** 10.1101/2023.10.07.23296007

**Authors:** Alexander D’Amico, Heejong Sung, Alejandro Arbona-Lampaya, Ally Freifeld, Katie Hosey, Joshua Garcia, Ley Lacbawan, Emily Besançon, Layla Kassem, Nirmala Akula, Emma E. M. Knowles, Dwight Dickinson, Francis J. McMahon

**Affiliations:** Intramural Research Program, National Institute of Mental Health, NIH, DHHS; Harvard Medical School

**Keywords:** Bipolar, cognition, genetics, heritability, SOLAR, Amish

## Abstract

**Background:** The basis of cognitive deficits in people with bipolar disorder (BD) has not been elucidated. These deficits may be the result of the illness or its treatment, but could also reflect genetic risk factors that are shared between BD and cognition. We investigated this question using empirical genetic relationships within a sample of patients with BD and their unaffected relatives.

**Methods:** Participants with bipolar I, II, or schizoaffective disorder (“narrow” BD, n=69), related mood disorders (“broad” BD, n=135), and their clinically unaffected relatives (n=227) completed tests of matrix reasoning, trail making, digit-symbol coding, semantic short-term recall, and affect recognition (DANVA2). General cognitive function (*g*) was quantified via principal components analysis (PCA). Heritability and genetic correlations were estimated with SOLAR-Eclipse.

**Results:** Participants with a “narrow” or “broad” BD diagnosis showed deficits in g, though affect recognition was not impaired. Cognitive performance was significantly heritable (*h^2^* = 0.322 for g, p<0.005). Coheritability between BD and cognition was small (coheritability for g & Narrow = 0.0184, for g & Broad = 0.0327) and healthy relatives of those with BD were cognitively unimpaired.

**Conclusions:** In this family sample, cognitive deficits were present in participants with BD, but social cognition, measured by affect recognition, was not impaired. Deficits were largely not explained by overlapping genetic determinants of mood and cognition. These findings support the view that cognition comprises separable social and non-social domains and that cognitive deficits in BD are largely the result of the illness or its treatment.

## Introduction

### Role of Cognition in Psychiatric Disorders

Cognitive impairment is a common feature of many psychiatric illnesses, including bipolar disorder (BD), and has been suggested as a transdiagnostic feature of psychiatric disorders (Sha et al. 2019). The relationship between cognition and psychopathology is complex and incompletely understood, but shared patterns of altered connectivity within and between cognitive networks have been identified across psychiatric disorders (Sha et al. 2019). Patients with BD display deficits in cognition during both affective episodes and periods of apparent euthymia. Unaffected relatives of BD patients have been suggested to display some deficits as well, although the data are inconsistent (Cardenas et al. 2016). Deficits in BD patients have been found in verbal learning and memory, executive functioning, response inhibition, attention/working memory, set-shifting, and processing speed (Cardenas et al. 2016). Previous studies suggest that common genetic factors influence cognition and schizophrenia risk (Lencz et al 2014), but it remains unknown whether cognitive deficits in BD are a result of the illness or its treatment, or reflect genetic risk factors that contribute to both BD and cognition.

### Bipolar Genetic Risk Spectrum

Bipolar disorder (BD) is one of the most common psychiatric illnesses, and is estimated to affect 1-4% of individuals (Merikangas et al. 2011). BD displays a wide range of symptoms, with the key defining feature being the cycling of mood and behavior between periods of depression, mania, and euthymia (Gordovez & McMahon 2020). Risk for BD has a strong genetic component. Studies comparing monozygotic and dizygotic twins have produced heritability estimates near 70% (Craddock & Jones 1999), while studies based on empirical relatedness have generated heritability estimates in the 25-45% range (Hou et al. 2016, Stahl et al. 2019). This “heritability gap” is a subject of ongoing investigation, but it is clear that relatives of those with BD are at increased risk for mental illness, especially mood and psychotic disorders.

Bipolar disorder also shares clinical features with MDD and SCZ. Depressive symptoms are a key feature of BD, and psychotic symptoms are not uncommon, especially during manic episodes. A variety of cognitive impairments have also been reported both during and between mood episodes (Cardenas & McMahon). These include deficits in attention, processing speed, verbal memory, and verbal fluency. However, it is unknown to what degree cognitive symptoms in BD are a core feature of the illness, as in SCZ, since cognitive impairments could be the result of BD or its treatment. One way to address this question is through study of “high risk” close relatives of people with BD. These relatives share genetic and familial risk factors, but do not have BD themselves. Such samples can be difficult to find and few have been published (Balanzá-Martinez et al. 2008). Extended families provide an alternative approach, where it is possible to collect more relatives and estimate their genetic risk in the absence of confounding due to illness or its treatment (Glahn et al. 2019).

The genetic architecture of BD is highly polygenic, including both common and rare variants. Many risk loci have been identified through Genome-Wide Association Studies (GWAS), the latest of which identified 64 independent risk loci. While these loci in aggregate appear to account for roughly 25% of genetic risk for BD, individually they contribute little to risk, and in most cases have not been parsed for specific risk genes (Gordovez & McMahon 2020). Genetic overlap between common genetic risk factors for BD and cognition appears small, in contrast to SCZ (Mullins 2021).

Rare variants may also play a role in risk for BD. Of particular interest are copy number variants (CNVs), small (∼30-1000 kb) stretches of DNA prone to duplication and deletion in the genome. CNVs have been heavily implicated in neurodevelopmental disorders (Kirov et al. 2014, Leppa et al. 2016, Sanders et al. 2011;70:863–85, Williams et al. 2010, Olsen et al. 2018, Gilissen et al. 2014, Pinto et al. 2010), schizophrenia (Kirov et al. 2014, Rippey et al. 2013, Szatkiewicz et al. 2014, Yuan et al. 2017, Bassett et al. 2008, Gulsuner et al. 2015, Ingason et al. 2011, Ahn et al. 2016), intellectual disability (Chiurazzi et al. 2020), and general cognition (Alexander-Bloch et al 2022). A duplication CNV on chromosome 16p11.2 and a deletion CNV on 3q29 have been most strongly implicated in risk for BD (Green et al. 2015). With the advent of whole genome sequencing (WGS) technologies, many more CNVs are being identified in the human genome, and may prove relevant to BD (Gordovez & McMahon 2020).

### Neurocognitive Measures in the Amish-Mennonite Bipolar Genetics Cohort

The Amish-Mennonite Bipolar Genetics (AMBiGen) cohort is composed of adult (≥18 yr) subjects with BD and related conditions, and their relatives. Most of the participants are part of genetically-isolated Anabaptist Amish and Mennonite communities with large families, uniform educational opportunities, and low rates of substance abuse. These communities are thus well-suited for the study of a range of genetically complex traits.

Here, we present the results of neurocognitive testing conducted on 373 AMBiGen participants. We examine the heritability of various cognitive performance measures in those with and without a psychiatric diagnosis, explore relationships between cognitive performance, psychopathology, and CNVs, and quantify genetic overlaps (co-heritability) between cognition and mood disorders.

## Methods and Materials

### Participant Selection

Participants in the AMBiGen cohort, from locations across the United States, Canada, and Brazil, were offered the opportunity to participate in cognitive testing. Participants included probands with bipolar disorder I or II or schizoaffective disorder, as well as their blood relatives. All participants were 18 or more years of age, and were free of physical and neurological disorders and substance abuse.

### Diagnostic Assignments

Participants whose screening suggested a psychiatric disorder were administered the Diagnostic Interview for Genetic Studies (DIGS) (Nurnberger et al. 1994). DIGS results, along with medical records and the companion Family Interview for Genetic Studies (FIGS, NIMH Genetics Initiative 1992) were reviewed by clinicians, and DSM-V diagnostic codes were assigned to each participant.

Participants were assigned an “affected” or “unaffected” status based on diagnosis, according to two separate categorical systems. The “narrow” affected category included subjects with bipolar disorder I or II or schizoaffective disorder (n=69). The “broad” affected category (n=135) included major mood disorders such as depressive disorders, which were present in some relatives, as well as all “narrow” affected subjects. 227 participants were unaffected, and 11 were undiagnosed due to incomplete diagnostic information.

### Cognitive Testing

Cognitive tests were administered for each participant as shown in Table 1.

**Table 1.** Cognitive tests performed.

| Abbreviation | Test Name(s) | Cognitive Domain Tested |
| --- | --- | --- |
| Matrices | Wechsler Abbreviated Scale of Intelligence Matrix Reasoning Test | Spatial reasoning (Stano 2004) |
| Trails A | Trail Making Test A | Visual-motor processing speed (Bowie & Harvey 2006) |
| Digit | Digit Symbol Substitution Test | Processing speed (Jaeger 2018) |
| VLT | California Verbal Learning Test (North America) or Rey Auditory Verbal Learning Test (Brazil) | Verbal short-term memory (Beier et al. 2019) |
| DANVA2 | Diagnostic Analysis of Nonverbal Accuracy | Visual affect recognition (Nowicki & Duke 2008) |

### Genomic Analyses

SNP array genomic data were obtained for each participant from blood samples. Using proportions of shared SNPs, a pairwise empirical genetic relatedness matrix was constructed using KING (Manichaikul et al. 2010) for use in heritability estimation. KING accurately estimates genetic relatedness from first-degree to 3rd-degree relatives.

CNVs were called from whole exome sequencing data, via Copy number estimation using Lattice-Aligned Mixture Models (CLAMMS) (Packer et al. 2016). CLAMMS has good sensitivity and specificity in calling CNVs across a broad size range, but since it depends on coding variants CLAMMS does not precisely define breakpoints that fall in intergenic regions.

### Data Analysis

R statistical computing version 4.2.3 was used for data analysis (R core team 2023). The Grubbs test (Grubbs 1969) was used to identify and remove outliers in the cognitive test scores. Scores on each cognitive test were regressed against age and years of education, separately for North American and Brazilian subjects. Z-scores of residuals from these regressions were pooled across the two regions, and used in subsequent analyses. Quantitative variables were compared using linear regressions. Diagnostic groups (narrow and broad affected vs unaffected) were compared using Student’s t-Test, unless otherwise specified.

Although the degree of specificity with which domains of cognition should be regarded is a subject of long-standing debate (Dickinson et al. 2008), the general construct g is robust and well-established (Jensen 2002). The g factor is calculated as the first principal component of measures of cognitive ability, and has an estimated heritability of about 50%, based on twin study data (Polderman et al. 2015). A principal components analysis was conducted on scores on all cognitive tests excluding DANVA2, and principal component (PC) 1 was recorded as “*g*” for each subject. Another principal components analysis was conducted on scores on all cognitive tests including DANVA2, and PC1 was recorded as the global intelligence score “*gwd*”. For comparing cognitive measures with age, residual Z-scores were re-derived, omitting age from the initial regression.

SOLAR-Eclipse version 9.0.0 (SOLAR Eclipse) was used to perform heritability analyses with the empirical genetic relatedness matrix. Heritability values for each cognitive measure were calculated according to Equation 1, where h2 = heritability, σ^2^_G_ = additive genetic variance, and σ^2^_P_ = additive phenotypic variance. Coheritability values for each cognitive measure with the narrow and broad categories were calculated according to Equations 2 and 3, where h_xy_ = coheritability of x and y, ⍴G = genetic correlation.

For each unaffected participant, proximities to the narrow and broad categories were determined by recording the empirical relatedness value of his/her closest affected relative in the narrow and broad affected categories, respectively. General cognition compared to family average (compg) was calculated by averaging g values for all close relatives (participants with ≥ 0.4 empirical relatedness) to a given individual, and subtracting that average from the individual’s g value.

CNV risk scores were obtained as follows. Gene-level probability of haploinsufficiency (pHI) and triplosensitivity (pTS) scores (Collins et al. 2022), were summed across all genes with deletions and duplications, respectively, for each participant. These scores are measures of the deleterious effect of deletion or duplication, respectively, of a given gene. The maximum probability of loss of function intolerance (pLI) score (Karczewski et al. 2020) among deleted genes for each subject and among duplicated genes for each subject were recorded. The gene LCE3C was omitted from CNV analysis, as read alignment errors resulted in an inflated number of counts for its deletion and duplication.

## Results

### Descriptive Statistics

Out of 373 total participants, 222 (59.5%) lived in North America, and 151 (40.5%) lived in Brazil. 217 (58.2%) were female, and 156 were male (41.8%) (Figure S1). 135 (36.2%) participants fell into the broad “affected” diagnostic category, including 66 (17.7%) in the narrow “affected” category. 227 were “unaffected” (in neither narrow nor broad categories), and 11 (2.9%) were undiagnosed (Figure S2).

The relationship between age and cognitive functioning in healthy participants was examined using linear regressions (Figure 5). As expected, all cognitive measures correlated significantly with age, with the digit-symbol task showing the strongest correlation and the DANVA2 showing the weakest. On this basis, all scores were age-adjusted in subsequent analyses.

Linear regressions between all cognitive tasks revealed significant correlations (Table S1). DANVA2 showed the weakest phenotypic correlation with the other tasks (Table S1), but the phenotypic correlations were still significant (Table S2).

### Trait Heritability

Heritability estimates for all cognitive measures were calculated, both including and omitting the broad diagnostic category as a covariate; age, years of education, sex, geographic region, and narrow diagnostic category were also tested individually as covariates, but the broad diagnostic category was the only covariate to improve model fit (Figure 1). All cognitive measures were significantly heritable, except for the Matrices task, and the verbal learning task with the broad covariate. Heritability was greatest for the digit-symbol task, and smallest for the Matrices task.

**Figure 1.**
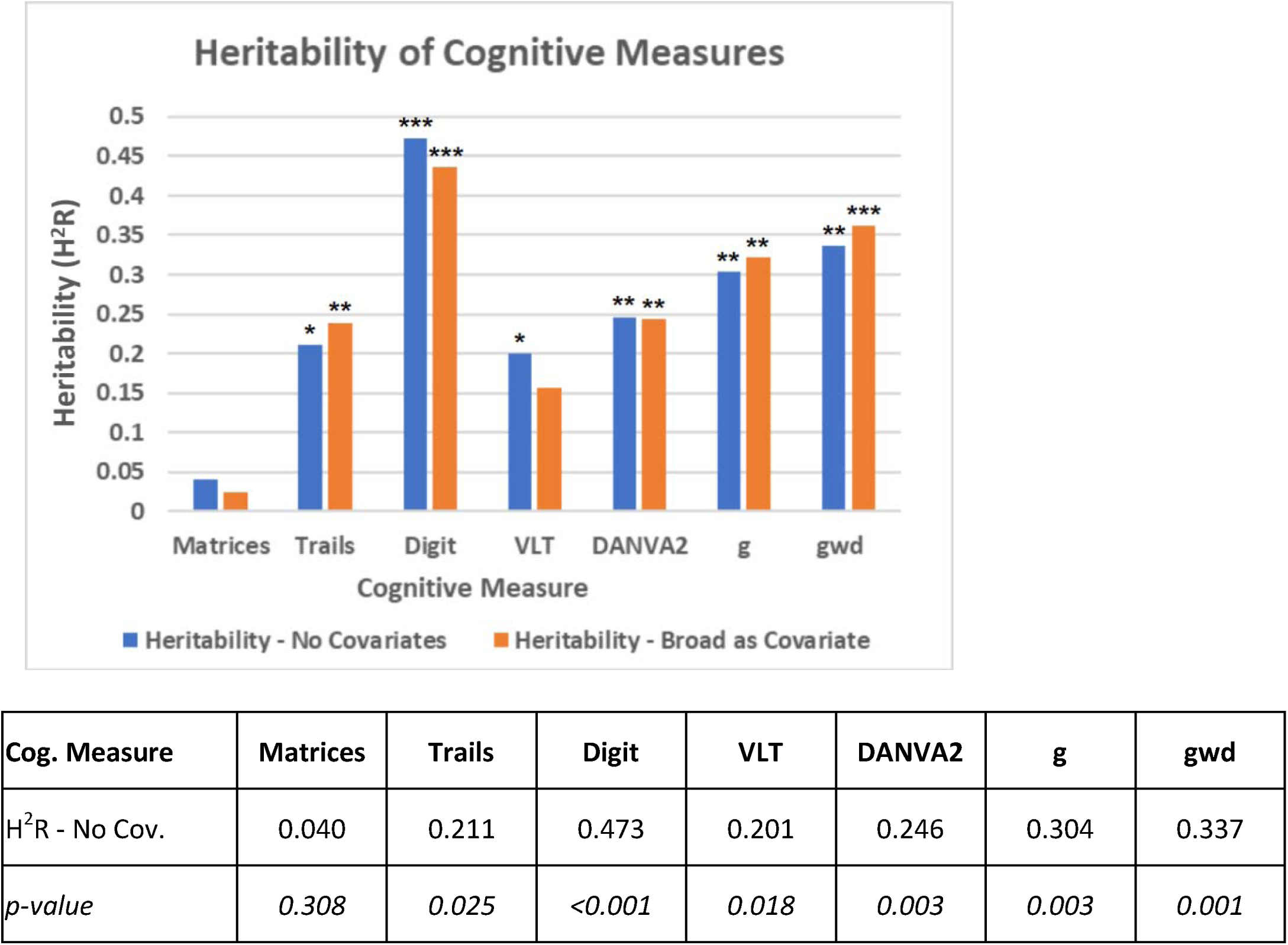
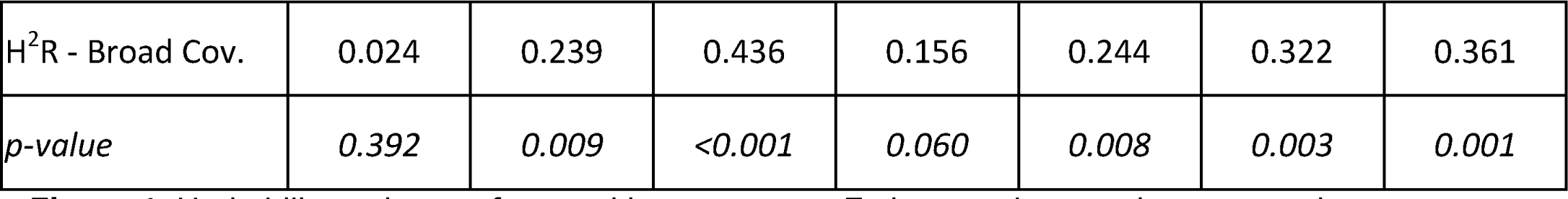
Heritability estimates for cognitive measures. Estimates shown using no covariates (blue) and with affection status (broad) as a covariate (orange). (*: p < .05, **: p < .01, ***: p < .001)

Estimates were also calculated for coheritability of each cognitive measure with each diagnostic group, using genetic correlation values according to Equation 3 (Figure 2). Many of the coheritability values were > 0.01, but no values were significant at the level of p < 0.05.

**Figure 2.**
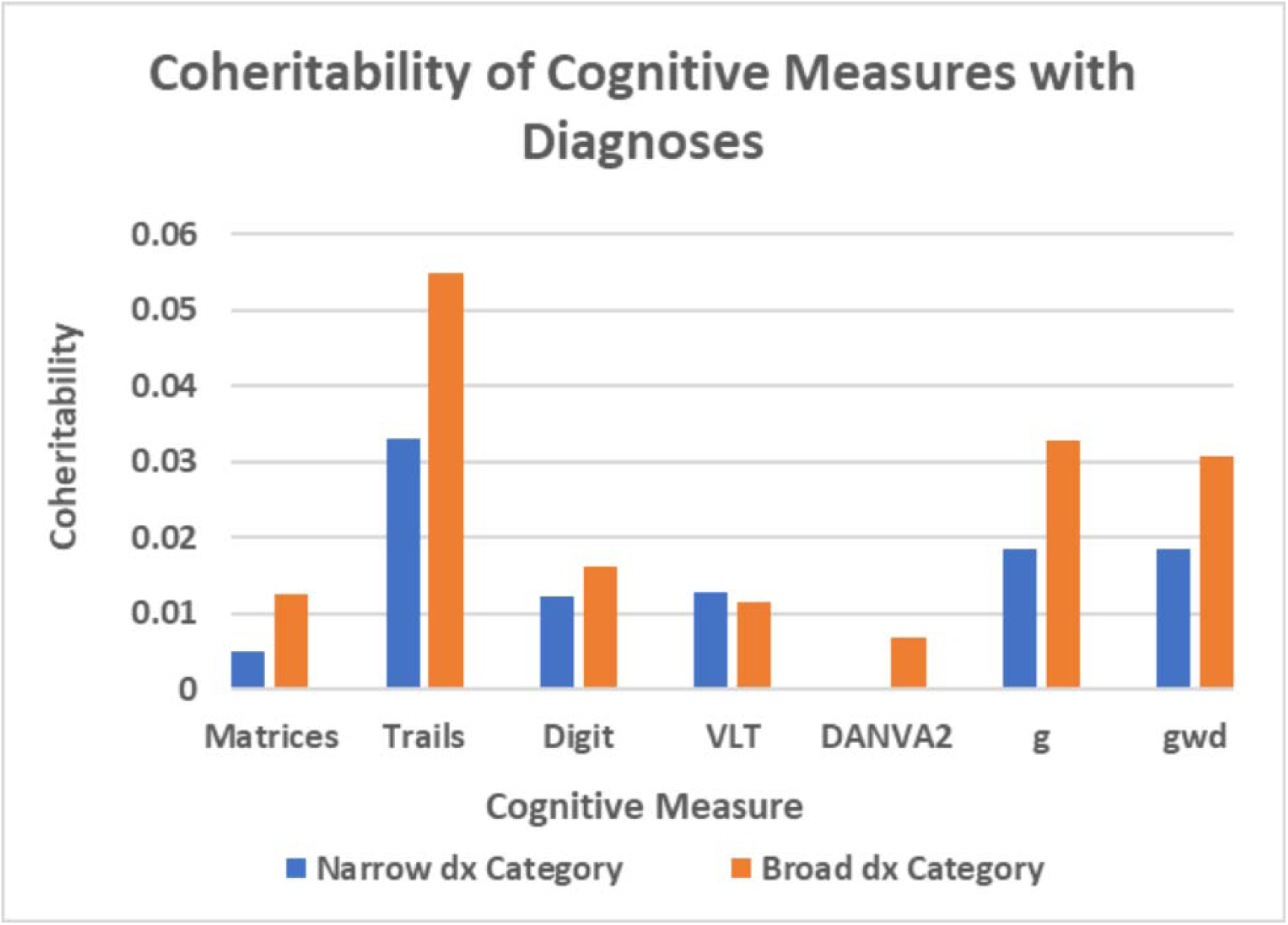
Co-heritability between cognitive measures and affection status according to narrow (blue) or broad (orange) diagnostic groups. All p > 0.05.

### Cognitive Deficits

Cognitive measures were compared between affected and unaffected participants, according to both narrow and broad diagnostic groups using Student’s t-test (Figure 3). General and global cognition scores were significantly different among affected participants in each diagnostic group. At the task level, the Trails A and Matrices task scores were significantly different for participants affected according to the broad definition; the Trails A task was significantly different for participants affected according to the narrow definition.

**Figure 3.**
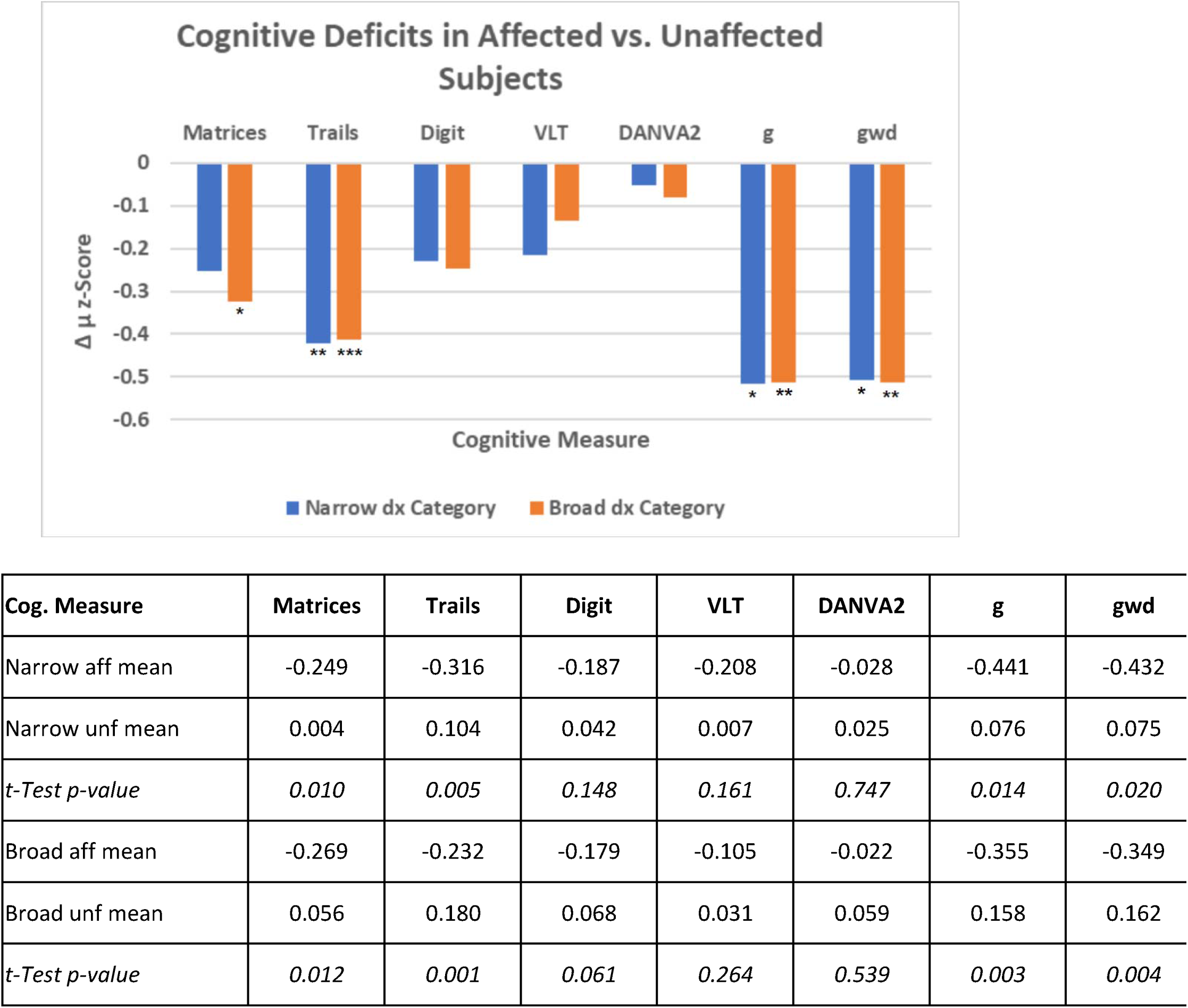
Differences in mean z-scores on cognitive tasks in affected (aff) and unaffected (unf) participants, according to both narrow (blue) and broad (orange) diagnostic groups. Welch Two Sample t-Test p-values are represented. (*: p < .05, **: p < .01, ***: p < .001)

Genetic proximity (proportion of SNPs shared with closest affected relative) values to both the narrow and broad diagnostic categories clustered near 0 and 0.5, and to a lesser extent 0.25 (Figures S11&12). These values represent pairs of individuals who are unrelated, first degree and second degree relatives respectively. The relationship between cognitive functioning in healthy participants and genetic proximity to an affected relative was examined using linear regressions of general cognition against genetic proximity to the most closely-related affected relative. No relationship was found under either the narrow or broad diagnostic groups (Figure 4). Linear regressions for all other cognitive measures vs proximity to each diagnosis group were all non-significant at p<0.2 (Table S3).

**Figure 4.**
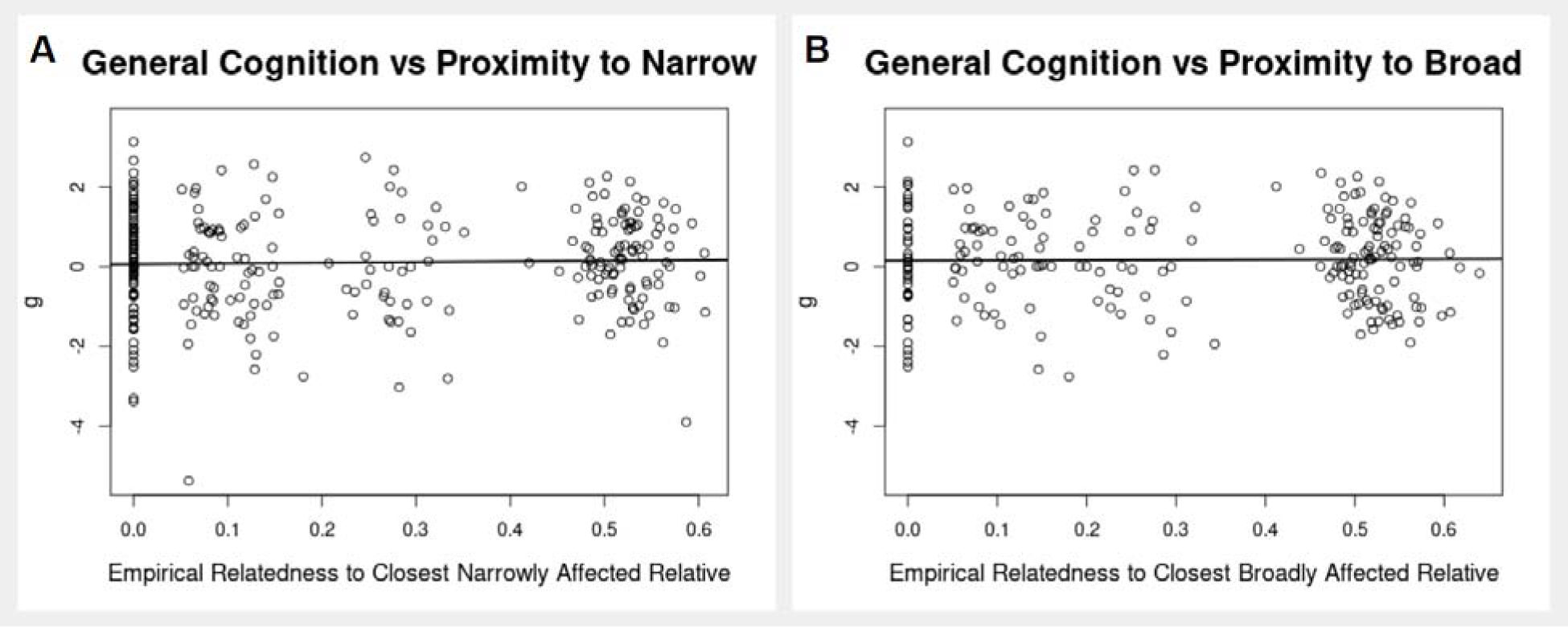
Scatter plots of general cognitive functioning vs. proximity to the “narrow” (**A**) and “broad” (**B**) diagnostic categories among unaffected subjects. Linear regressions results - **A**: R2 = 0.00095, p = 0.60; **B**: R2 = 0.00018, p = 0.84.

**Figure 5.**
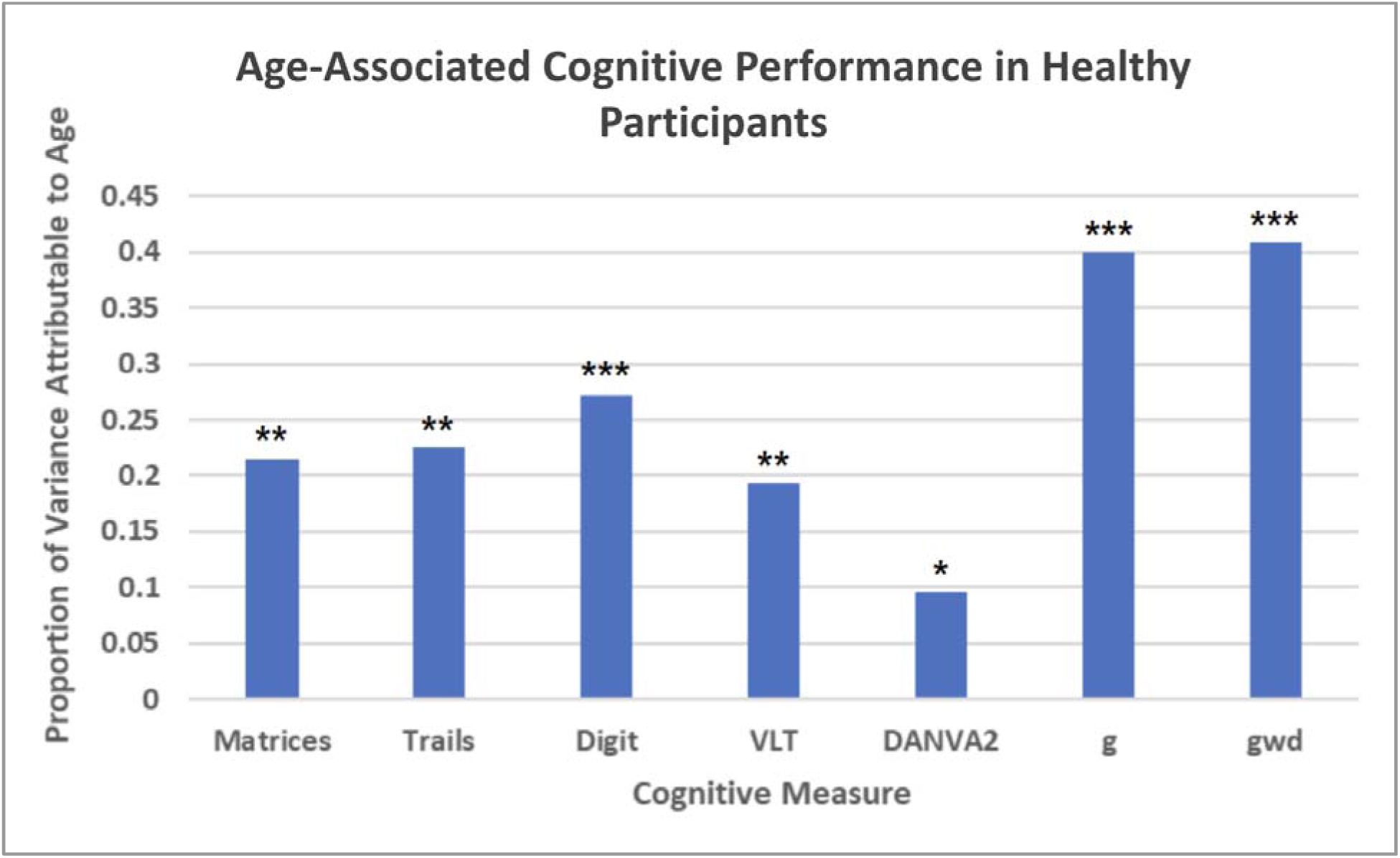
Adjusted R^2^ values are shown for linear regressions of each cognitive measure against participant age at testing. (*: p < .05, **: p < 10^-5^, ***: p < 10^-10^) Matrices Adj. R^2^ = 0.214, p = 1.17E-13. Trails Adj. R^2^ = 0.226, p = 2.02E-14. Digit Adj. R^2^ = 0.272, p < 2.20E-16. VLT Adj. R^2^ = 0.193, p = 2.45E-12. DANVA2 Adj. R^2^ = 0.096, p = 1.18E-06. g Adj. R^2^ = 0.400, p < 2.20E-16. gwd Adj. R^2^ = 0.408, p < 2.20E-16. Correlation direction was negative in all regressions.

Since genetic correlations between polygenic traits such as BD and cognition decay rapidly with each meiosis, we repeated this analysis after splitting the genetic proximity scores into three bins. This revealed significant genetic correlations between g and proximity to both narrow and broad cases only for proximity scores greater than 0.4 (LM g ∼ narrow proximity [where proximity > 0.4] adj. R^2^ = 0.03286, p =0.0417; LM g ∼ broad proximity [where proximity > 0.4] adj. R^2^ = 0.04665, p = 0.01484.

### Copy Number Variants

Among CNVs, copy numbers ranged from 0 to 6 (Figure S13). Sums of pHI and pTS scores per subject clustered around 0, with tails to the right (Figures S14&15). Maximum pLI score distributions in deleted and duplicated genes had peaks at the extremes of 0 and 1 (Figures S16&17).

The relationship between cognitive functioning and CNVs was examined using linear regressions of general cognition against sum of pHI and pTS scores (Figure S18). Neither score was significantly correlated with general cognition. Maximum pLI scores among deleted and duplicated genes for each participant were also regressed against g, and these, as well as pHI and pTS scores, were regressed against a family-standardized general cognition (compg) score (Table S4). Max deleted pLI showed a small positive correlation with g (Table S4), but this was not significant at p<0.05. Furthermore, mean g values in subjects with max deleted pLI < 0.1 were not significantly different from mean g values in subjects with max pLI> 0.9 (Table S5).

### Dimensions of Cognition

The degree to which the DANVA2 task measures a separate dimension of cognition from the other four tasks was examined by regressing DANVA2 scores against the first four principal components of the five tasks including DANVA2 (Figure 7). DANVA2 scores correlated more strongly with PC2 than with PC1 (Adj. R2 = 0.81 vs 0.21).

## Discussion

### Key Findings

As expected, performance on some cognitive tasks was impaired in affected participants, although affect recognition was not impaired in this sample. Also as expected, g and most individual cognitive tasks were significantly heritable, though heritability values varied between tasks. Only a small relationship was found between cognitive functioning and polygenic risk for BD, with similar estimates obtained from co-heritability and from genetic proximity analyses. Thus the cognitive impairments we observed among affected participants were largely unexplained by shared genetic determinants between mood disorder and cognition.

### This study has several important limitations

This cross-sectional study cannot distinguish between cognitive impairments that precede illness onset and those that follow. In order to further distinguish the cognitive effects of BD and/or its treatment from the effects of associated genes, longitudinal testing of individuals at risk for BD is needed. A decline in cognitive performance after illness onset would lend support to the hypothesis that cognitive deficits are a result of the illness or its treatment.

In regression models, t-tests and PCAs in this study, relatedness between individuals may serve as a confounding variable. In order to address this issue, future work examining the relationship between cognition and empirical relatedness to a BD patient among healthy subjects, who are themselves unrelated, is needed.

We did not detect an association between CNV burden and cognitive impairment in this sample, but this may reflect power limitations imposed by sample size and the relatively low rates of large deletions in this sample. Such deletions have the strongest impact on cognition (Stefansson et al 2014; Alexander-Bloch et al 2022) and are typically rare in BD samples (Charney et al 2019). Indeed, the few carriers of large deletions in the present sample had among the lowest cognitive performance scores. The lack of significant CNV effects does not appear to be driven by aberrant CNV risk scores, as risk score distributions followed expected patterns for pHI and pTS (Collins et al. 2022) (Figures S14&15), and for pLI (Karczewski et al. 2020) (Figures S16&17). The small positive correlation we detected between g and pLI scores of deleted genes (Table S4) is likely spurious, probably owing to a poor fit of the linear regression model we used to the (non-linear) distribution of the data (Table S5).

We have not investigated the potential impact of rare, damaging single nucleotide variants in this sample. Such variants are known to have impacts on psychopathology and cognition (Chen et al. 2023). However, if such variants contribute to cognitive impairments observed in this sample, the contribution must be small, since inherited variants would be shared by ∼50% of first-degree relatives, and they did not display any cognitive deficits.

### Trait Heritability

The heritability of general intelligence, “g” has been well documented, though estimates have clustered around 50% (Polderman et al. 2015), higher than the 30% estimated here (Figure 1). Differences in study samples and in the test batteries used to derive “g” might account for this variation. The limited battery of tests we employed is sensitive to variations in task-relevant skills and performance. The low heritability estimate for the Matrices task in this sample contrasts with prior literature (King et al. 2019) and may reflect special characteristics of our study sample.

### Cognitive Deficits in Affected Individuals

Lower cognitive scores among participants in both the narrow and broad diagnostic categories (Figure 3) support prior findings of cognitive deficits in bipolar disorder (reviewed in Cardenas et al. 2016). The 0.5 standard deviation decrease in g among cases is well within reported effect size ranges in previous studies (Bourne et al. 2013, Mann et al. 2011). In the present sample a decrease was observed across all tasks, but was disproportionately attributable to the Trails A task, which may be sensitive to slowed motor function.

Cognitive impairments and mood disorder coheritability estimates showed a similar pattern, with Trails A showing the greatest effects and DANVA2 showing the smallest effects (Figures 2&3). The low coheritability between cognitive functioning and mood disorder diagnosis in this sample suggests that if there is a genetic overlap between cognition and BD, it is much smaller than that observed in other psychiatric conditions such as schizophrenia (Lam et al. 2019) or psychosis (Knowles et al. 2021). However, the co-heritability analyses included some affected individuals, so cannot fully distinguish between the effects of shared genes and the effects of illness or its treatment.

To address this limitation, we tested for a correlation between cognition in participants with no psychiatric diagnosis and their genetic proximity to relatives with mood disorders. The complete lack of correlation (Figure 4, Table S3) suggests that the cognitive deficits we observed among affected participants do not reflect overlapping genetic determinants between mood disorders and cognition. This finding is supported by the polygenic risk score (PRS) literature, which shows no correlation between bipolar disorder PRS and cognitive functioning (Ranlund et al 2018, Ohi et al. 2022).

### Dimensions of Cognition

The multidimensional structure of cognitive ability has been a subject of ongoing debate in the cognitive psychology literature (Dickinson et al. 2008). The relative sparing of DANVA2 scores among affected participants (Figure 3) suggests that BD and other mood disorders may impact distinct dimensions of cognition to varying degrees. The DANVA2 measures affect recognition through photos of faces, so is a more social task than the others we used, and is typically excluded from measures of g. Some neuropsychiatric disorders have differential impacts on distinct aspects of cognitive function. For example, patients with William’s syndrome display deficits in general cognition, but spared or improved performance in social cognition (Gagliardi et al. 2006). The strong loading of DANVA2 scores on PC2 of all tests in the present study (Figure 7) suggests that DANVA2 scores represent a separable social dimension of cognition. As previously discussed, this dimension appears unimpaired in BD in this sample (Figure 3).

This contrasts with a previous report that found a deficit in the DANVA2 task in BD, but that result was non-significant when controlling for participant age and years of education (Lee et al. 2022). Other studies have also failed to find deficits in social cognition in BD (Miskowiak et al. 2019).

While DANVA2 scores appear to be spared in affected participants, the scores do appear to share significant genetic determinants with general cognition. This is indicated by coheritability values on par with those observed among the measures of general cognition we used (Table S2). A more complete study of social cognition in BD should use additional measures in more culturally diverse populations.

## Conclusions

In this family sample, cognitive deficits were present in participants with BD, but social cognition, measured by affect recognition, was not impaired. Deficits were largely not explained by overlapping genetic determinants of mood and cognition. These findings support the view that cognition comprises separable social and non-social domains and that cognitive deficits in BD are largely the result of the illness or its treatment.

## Data Availability

Data and associated biological specimens are available to qualified investigators through dbGAP (phs000899.v1.p1)

## Acknowledgements

Funded by the NIMH Intramural Research Program, grant 1ZIAMH002843 and protocol 80 M 0083. Human materials were collected by informed consent under protocol.

We thank the members of the NIMH Intramural Research Program and the Human Genetics Branch. We thank Ellen Leibenluft, MD, (NIMH) and Melissa Brotman, PhD, (NIMH) for assisting with selection of cognitive measures, as well as Aaron Alexander-Bloch, MD, PhD (UPenn. Dept. of Psychiatry) for assistance annotating CNVs. We thank Alan Shuldiner and the Regeneron Genetics Center for exome sequencing. We also thank all participants who donated blood and gave interviews for this project, as well as their families. Data and associated biological specimens are available to qualified investigators through dbGAP (phs000899.v1.p1)

## Supplemental Figures and Tables

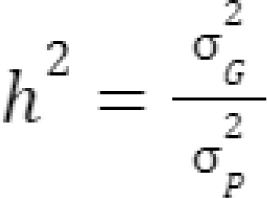

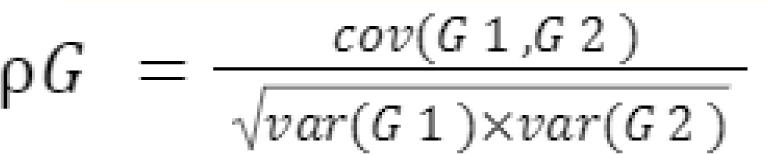

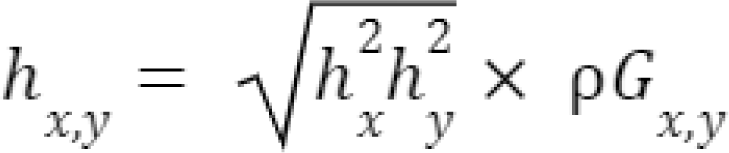

**Figure S1.**
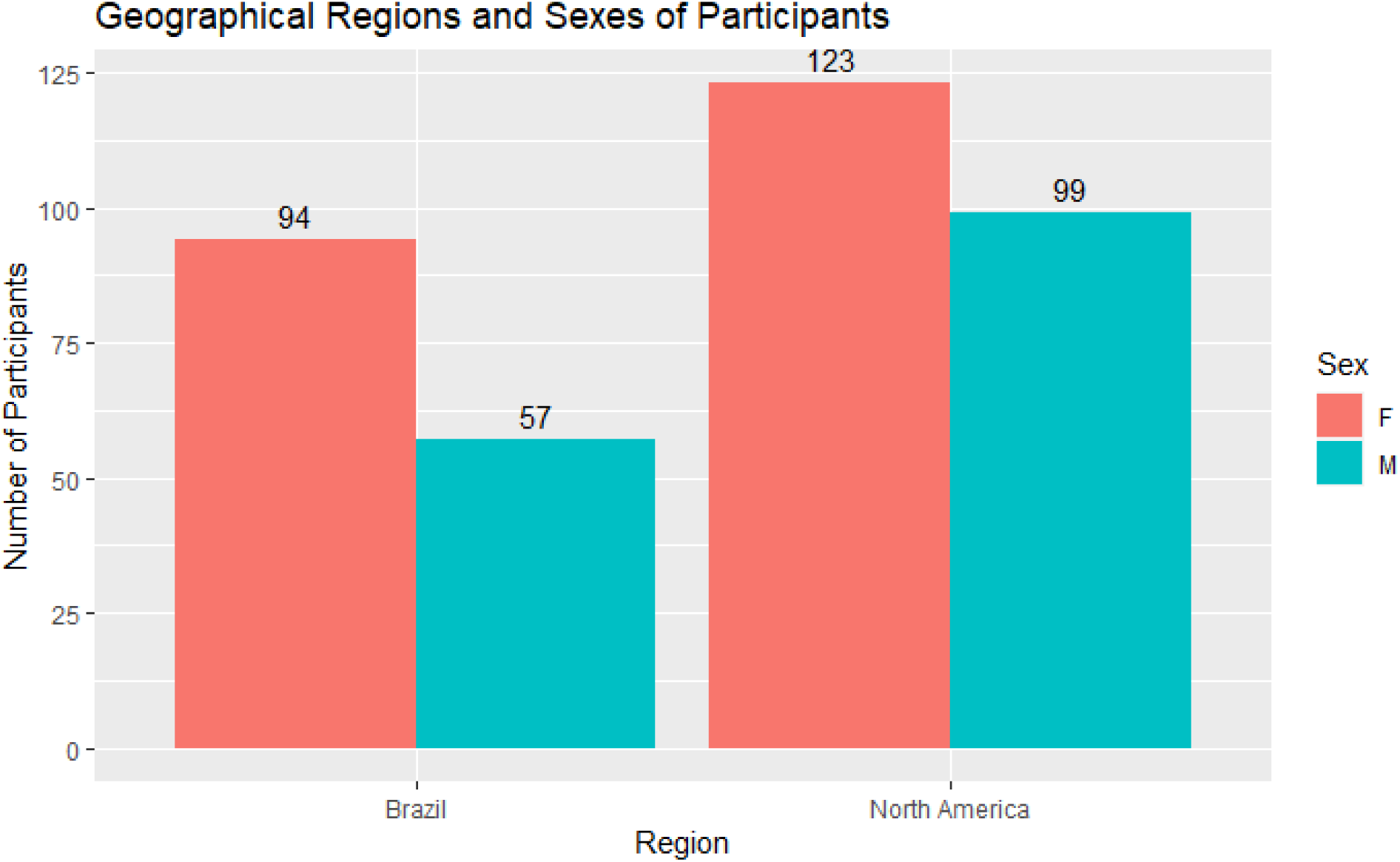
Distribution of study participants across sexes and geographical regions.

**Figure S2.**
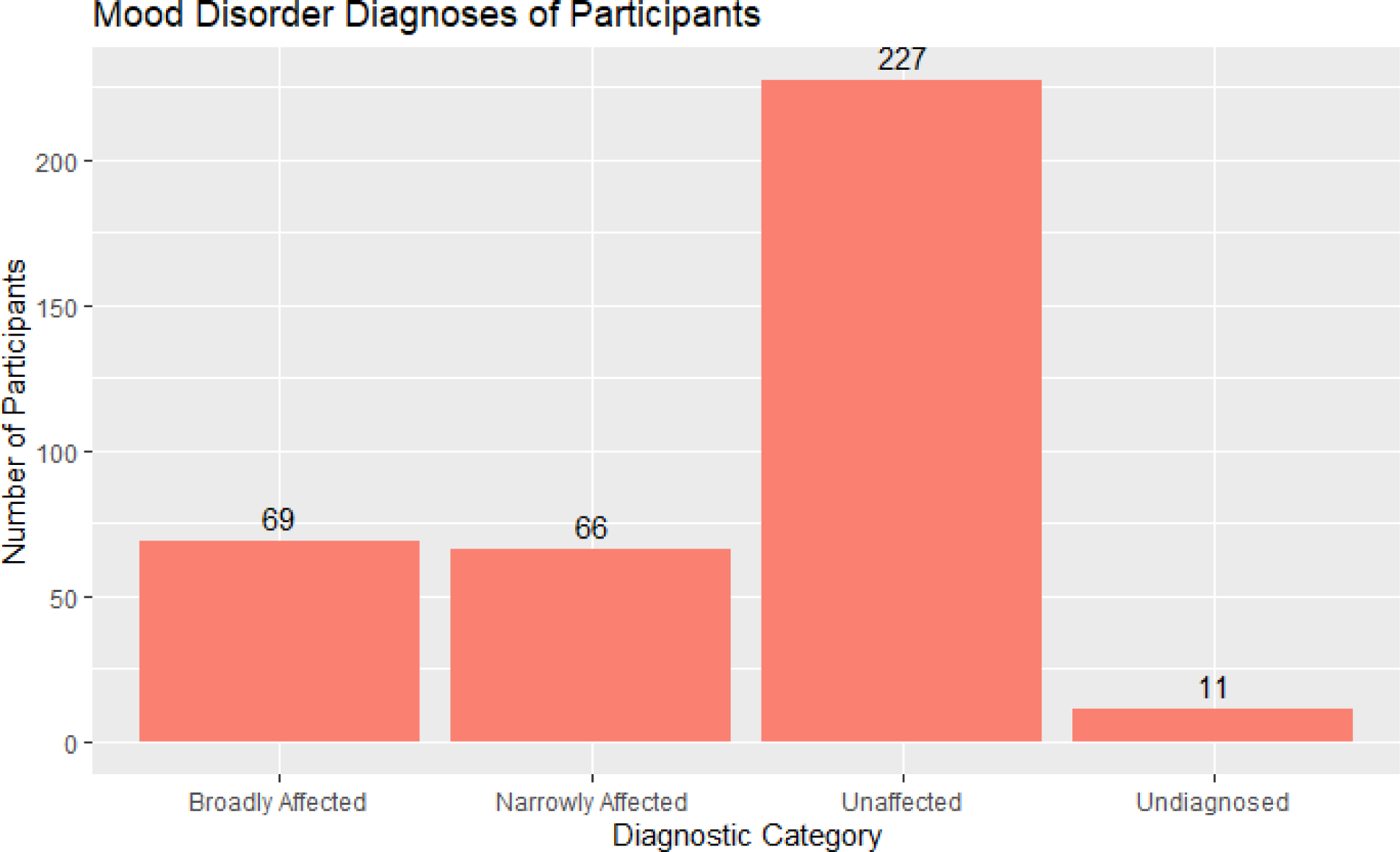
Distribution of study participants across diagnostic categories.

**Figure S3.**
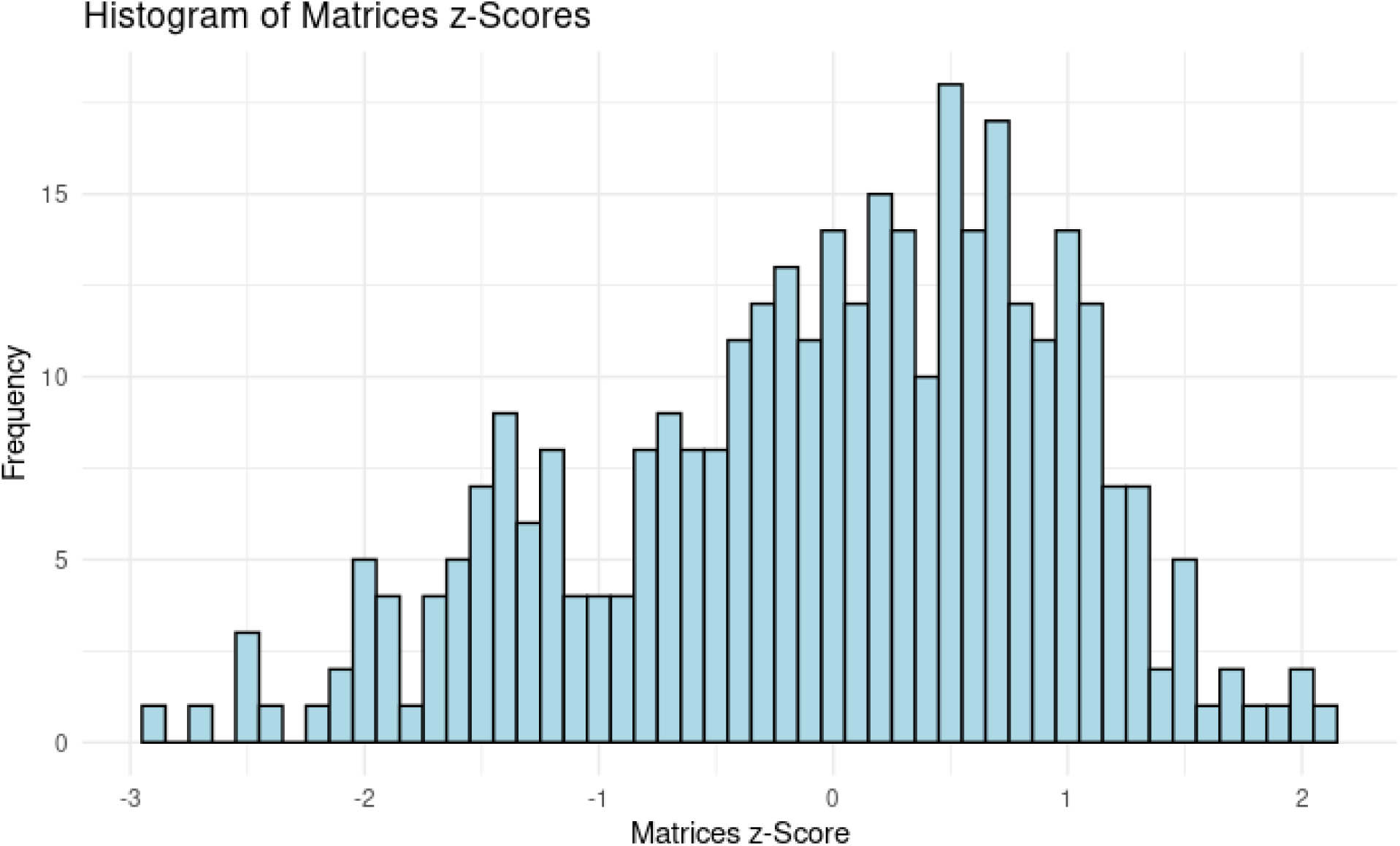
Distribution of study participant age and education-adjusted z-scores on the Wechsler Abbreviated Scale of Intelligence Matrix Reasoning Test (Matrices). µ = -0.034, SD = 0.99.

**Figure S4.**
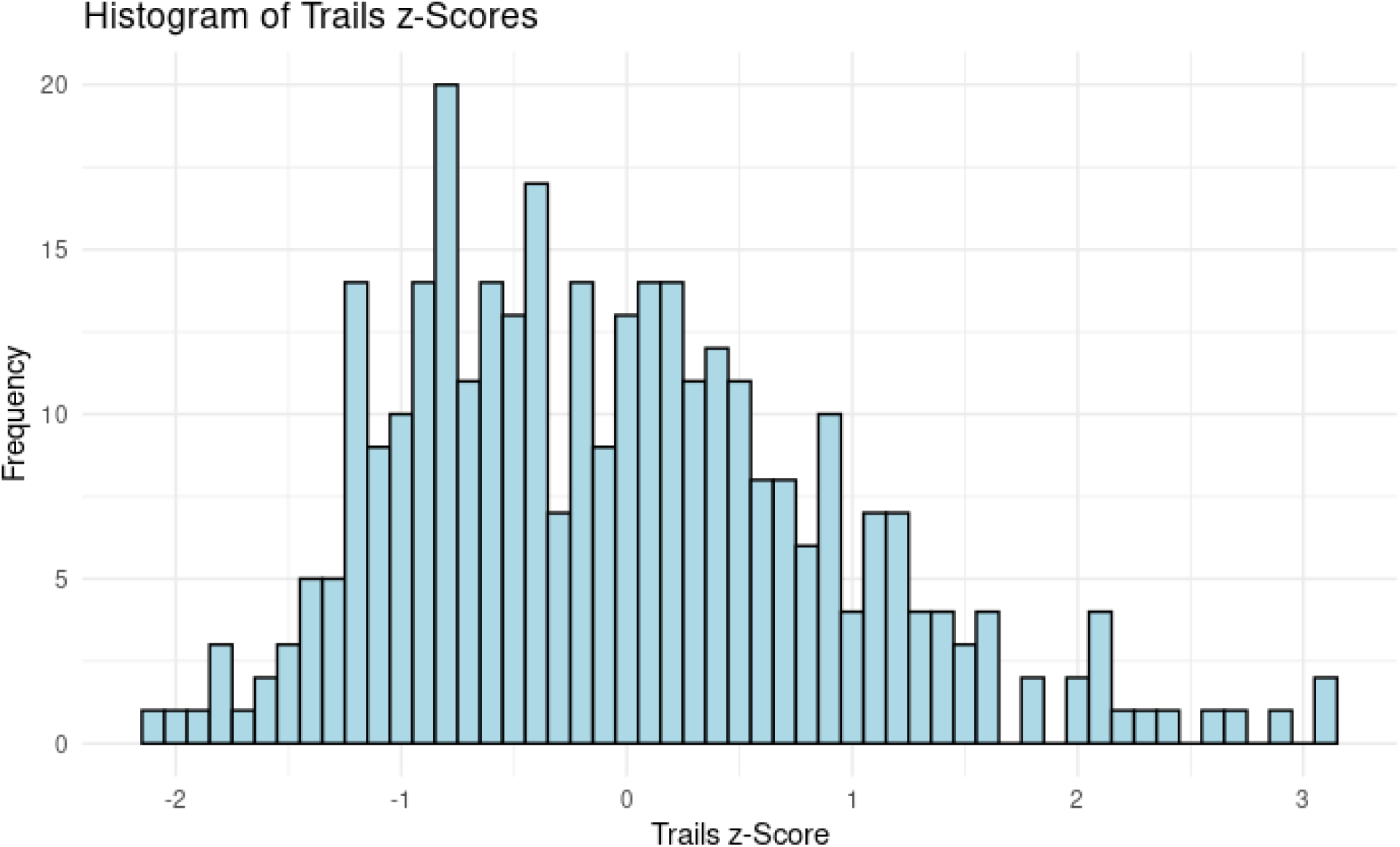
Distribution of study participant age and education-adjusted z-scores on the Trail Making Test (Trails). µ = -0.039, SD = 0.96.

**Figure S5.**
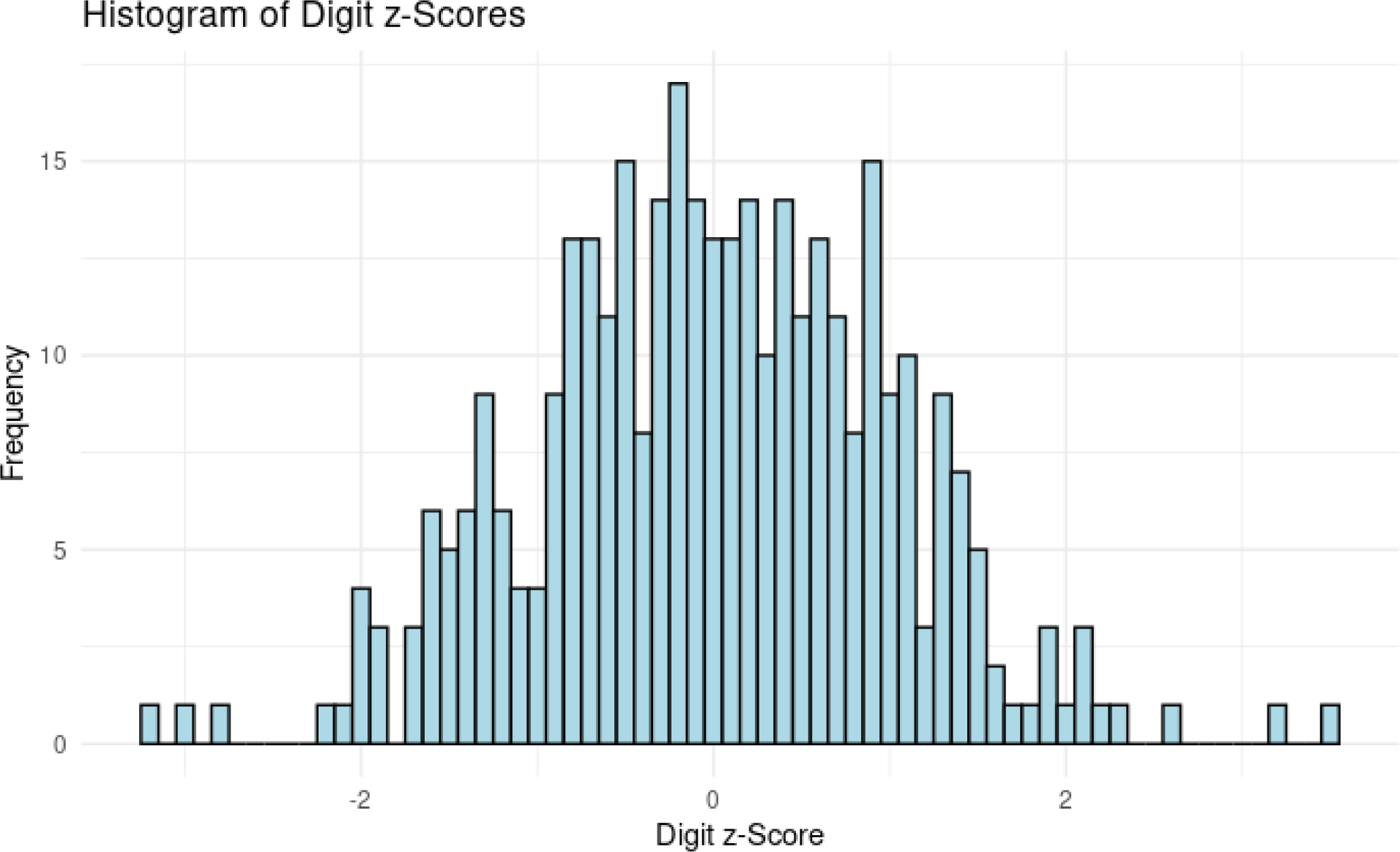
Distribution of study participant age and education-adjusted z-scores on the Digit Symbol Substitution Test (Digit). µ = 0.007, SD = 1.01.

**Figure S6.**
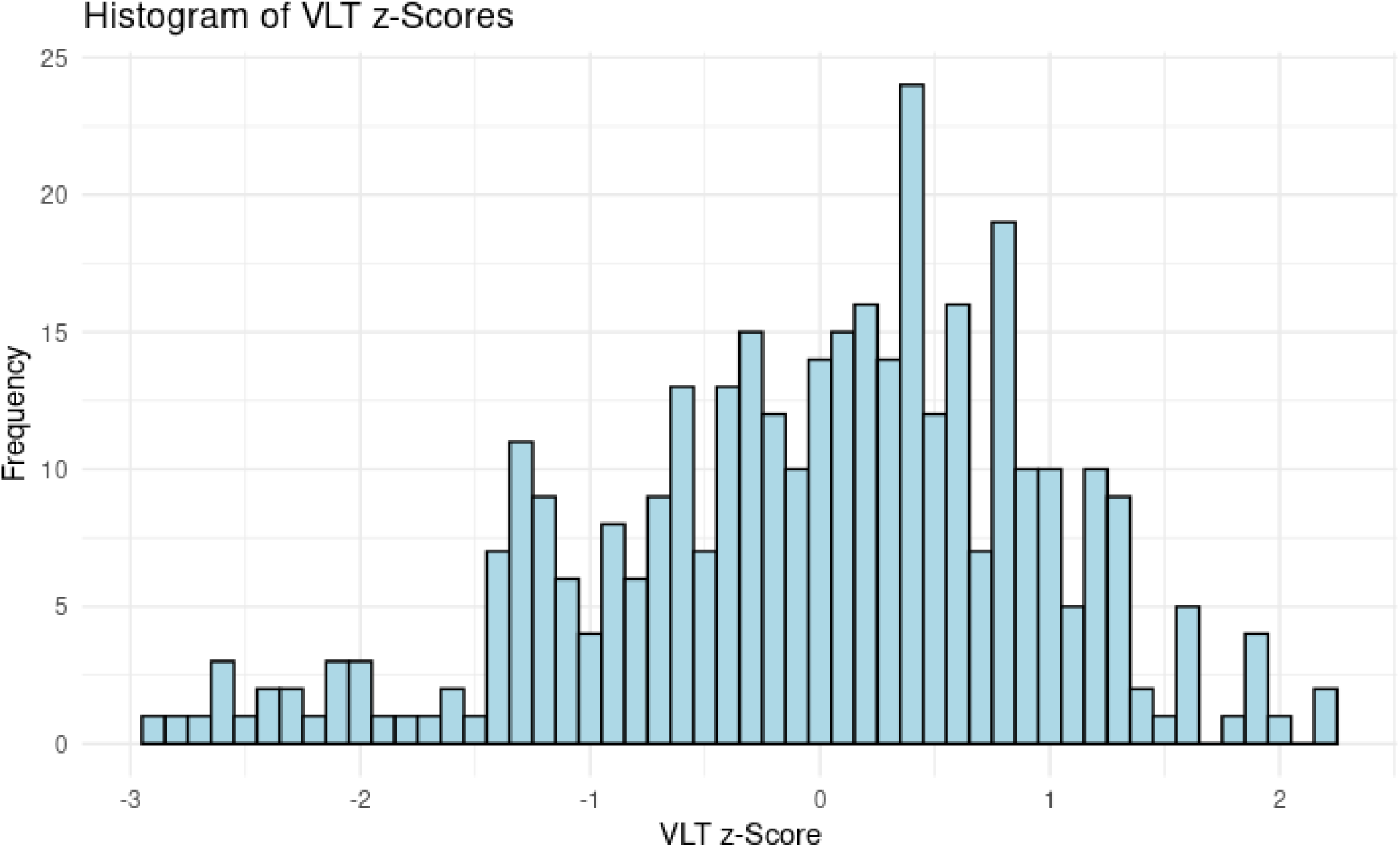
Distribution of study participant age and education-adjusted z-scores on the California Verbal Learning Test or Rey Auditory Verbal Learning Test (VLT). µ = -0.026, SD = 0.98.

**Figure S7.**
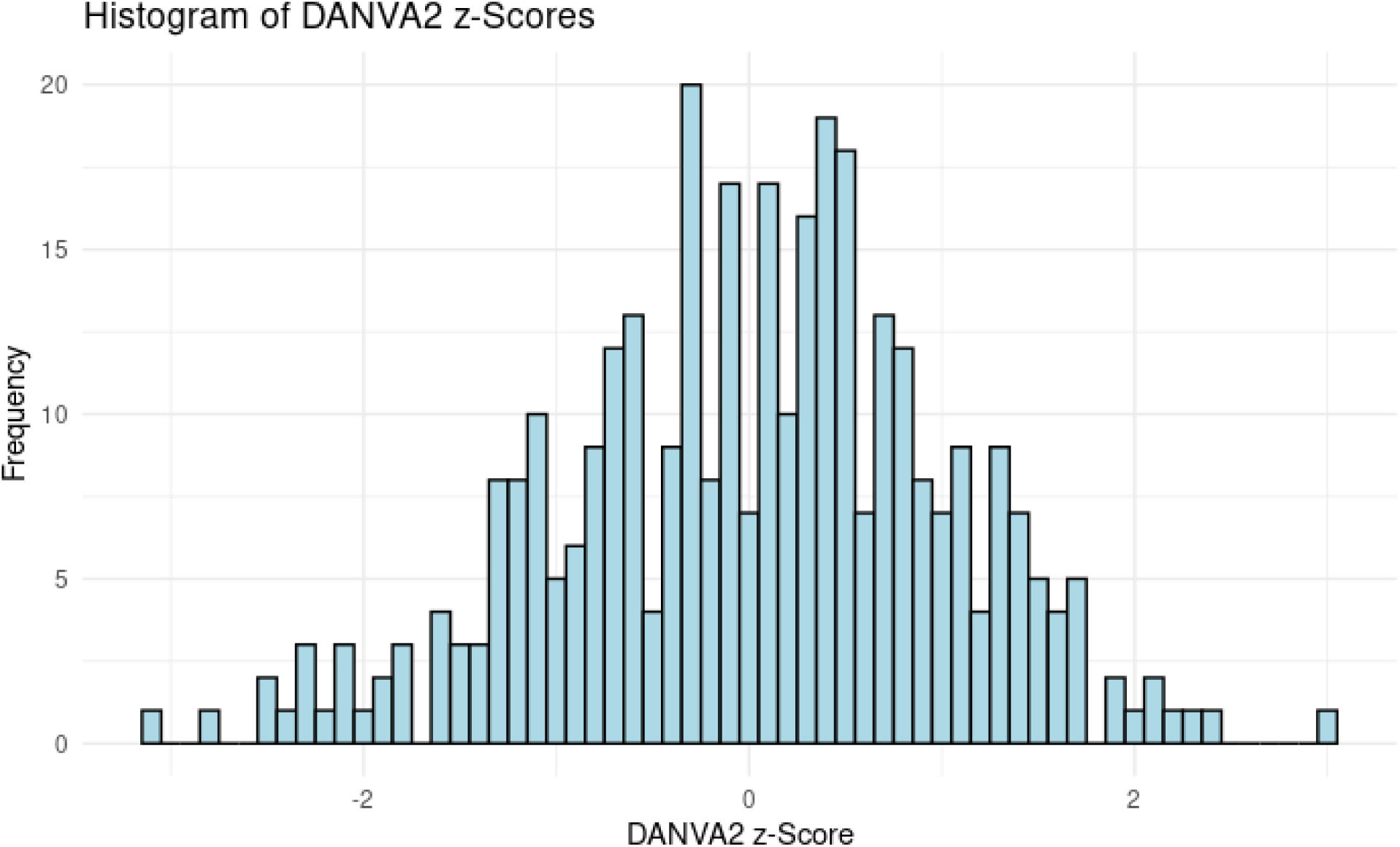
Distribution of study participant age and education-adjusted z-scores on the Diagnostic Analysis of Nonverbal Accuracy (DANVA2). µ = 0.017, SD = 1.01.

**Figure S8.**
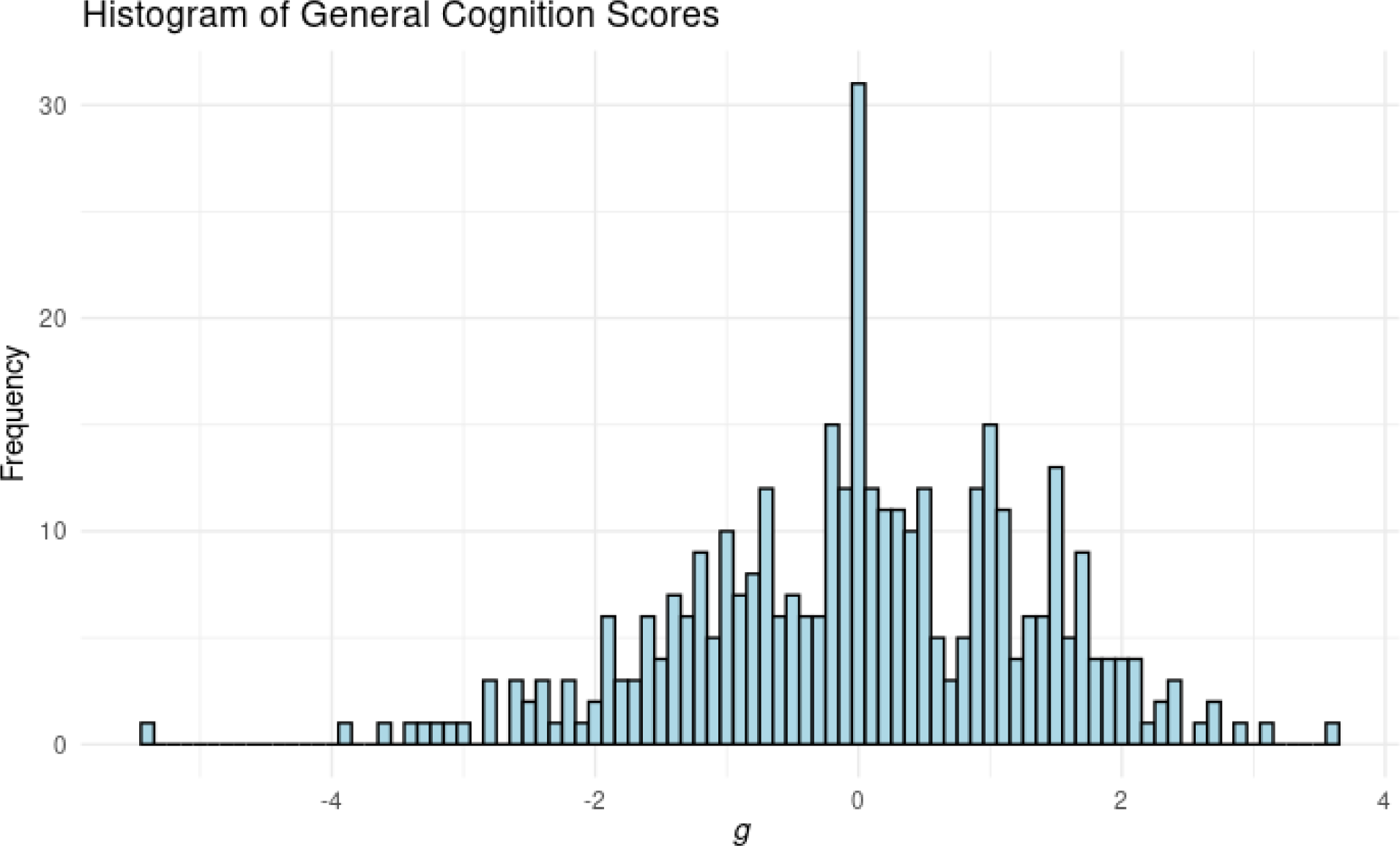
Distribution of study participant general cognition factor (*g*) scores. µ = 0, SD = 1.32.

**Figure S9.**
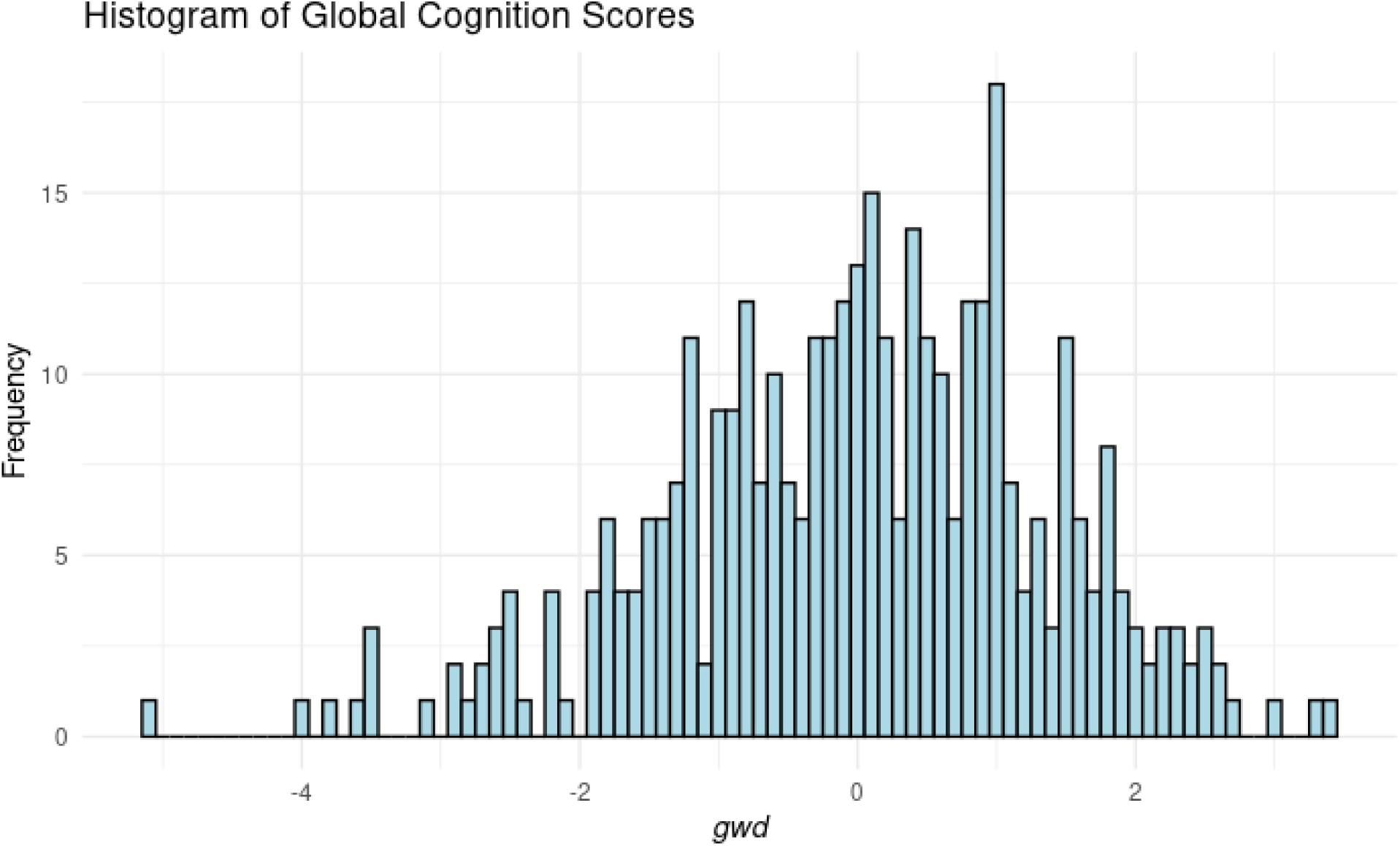
Distribution of study participant global cognition factor (*gwd*) scores. µ = 0, SD = 1.35.

**Figure S10.**
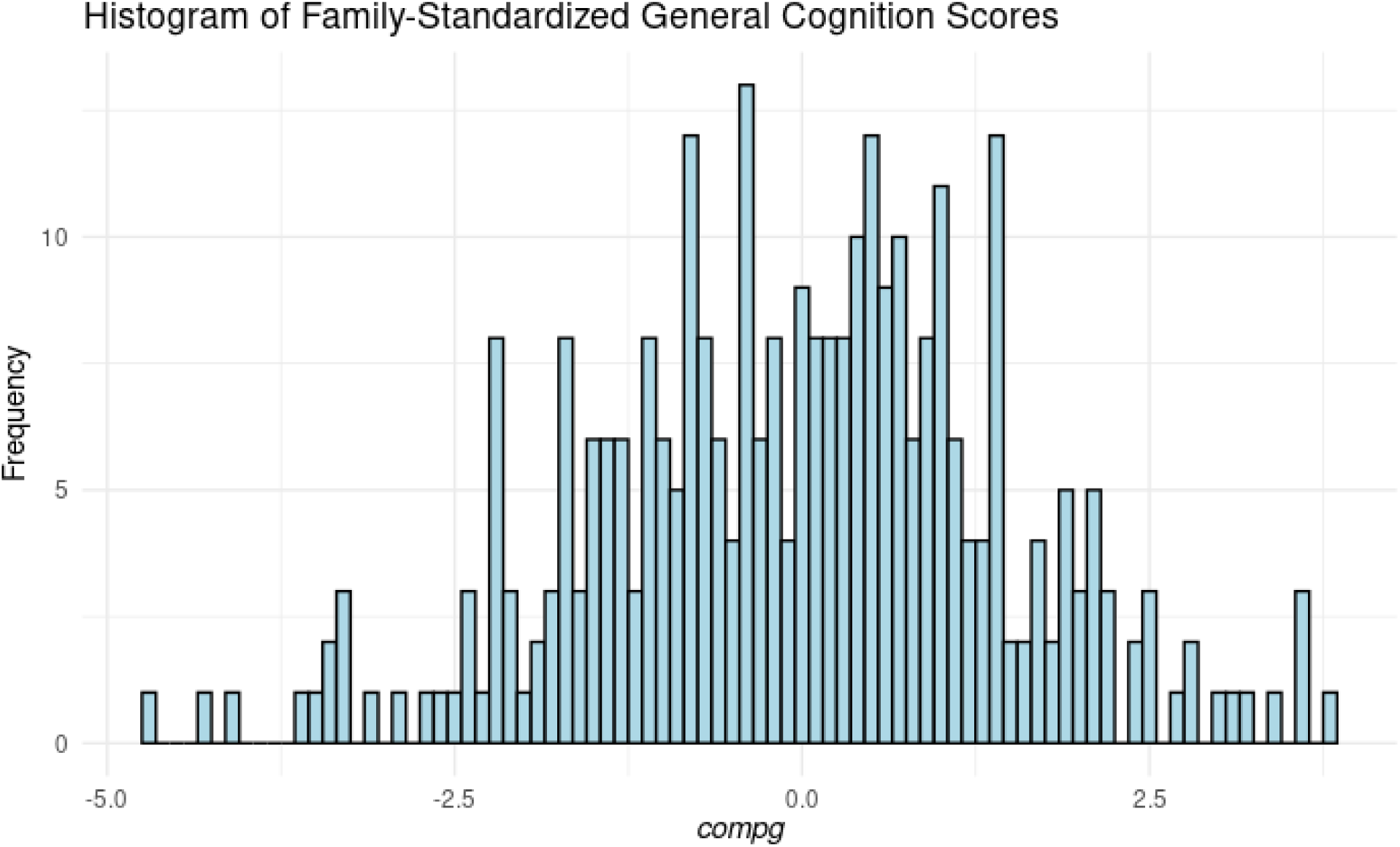
Distribution of study participant general cognition compared to family average (compg) scores. µ = 0, SD = 1.49.

**Figure S11.**
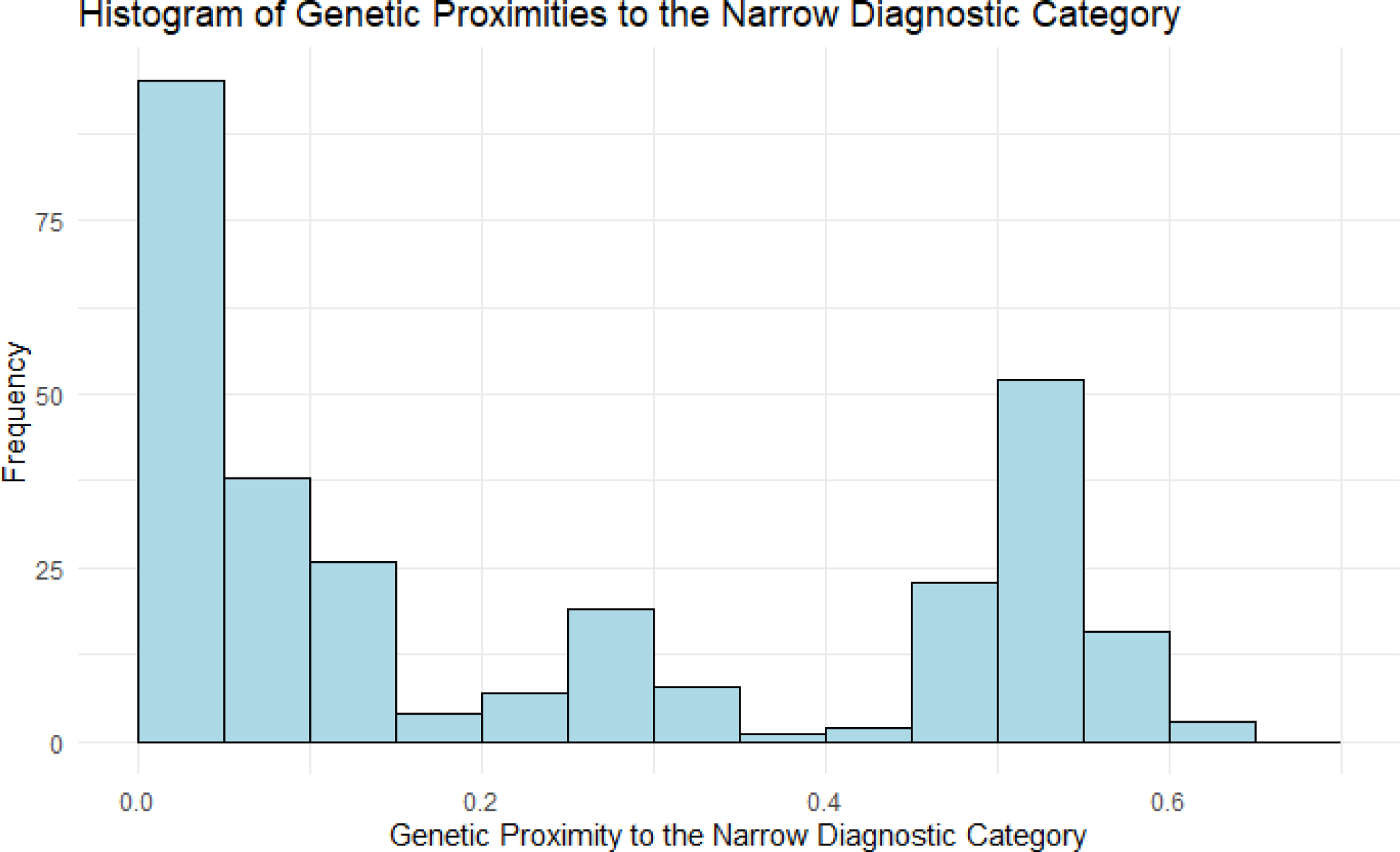
Distribution of unaffected study participant genetic proximities to the narrow diagnostic category.

**Figure S12.**
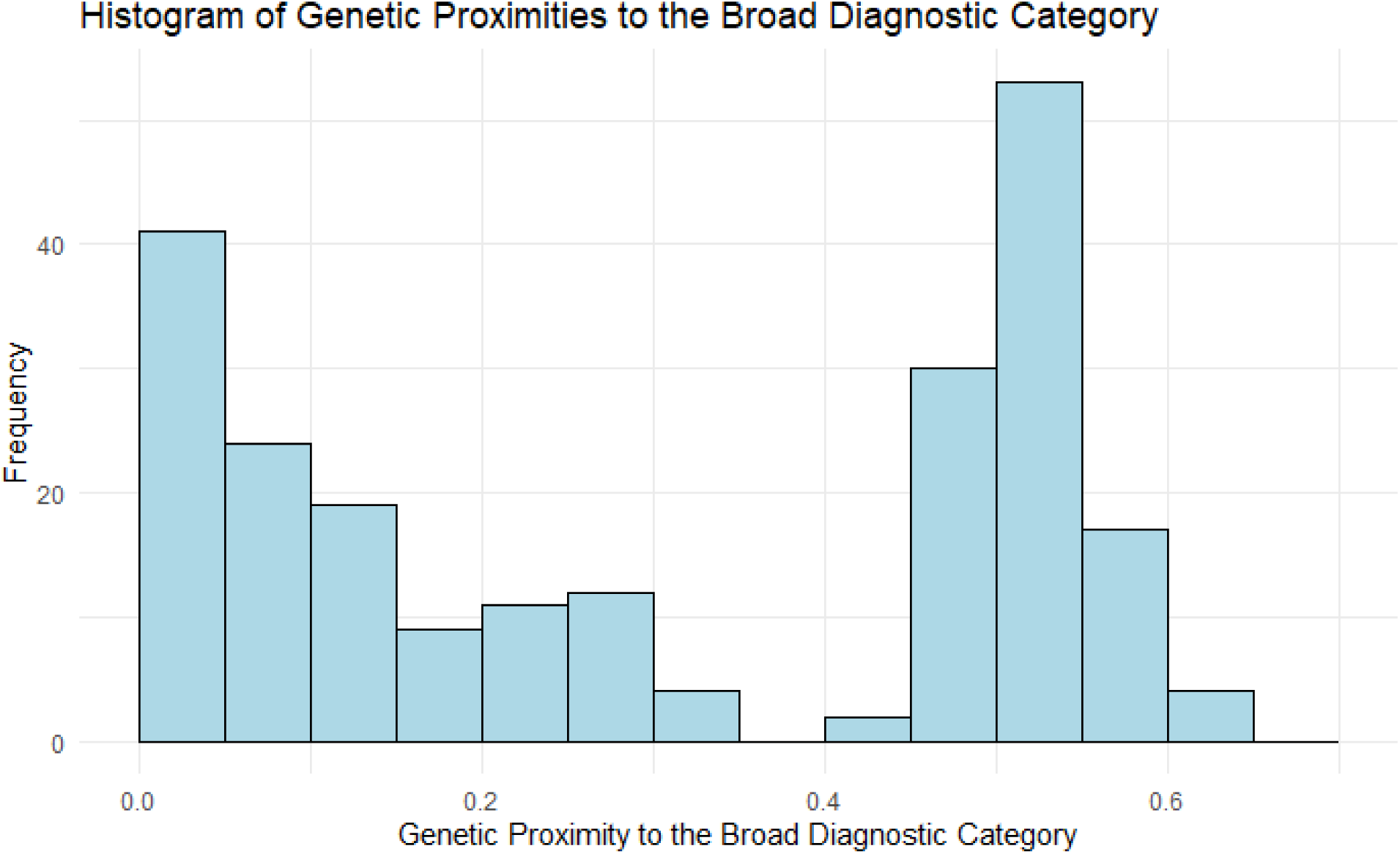
Distribution of unaffected study participant genetic proximities to the broad diagnostic category.

**Figure S13.**
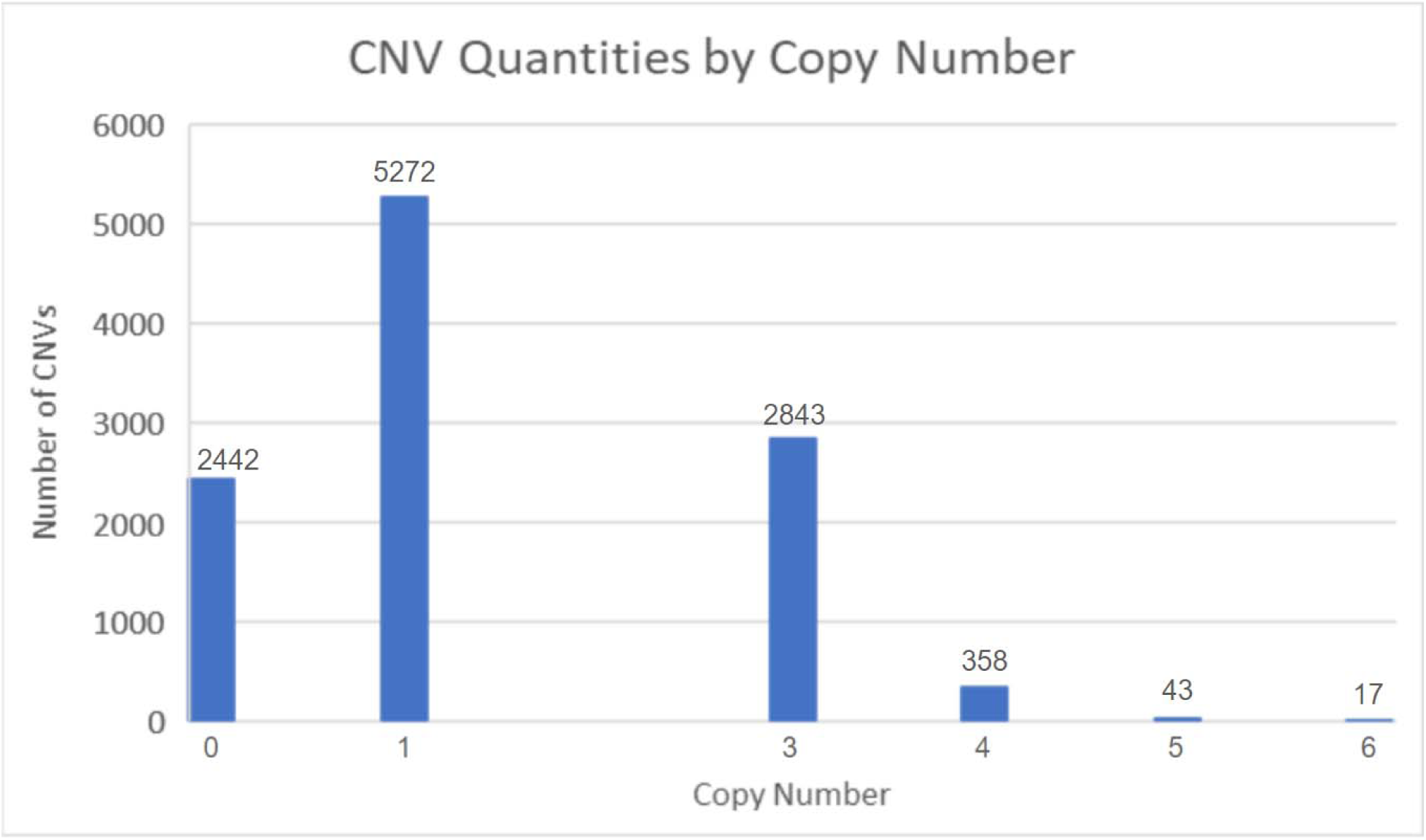
Distribution of copy numbers among duplicated and deleted copy number variants.

**Figure S14.**
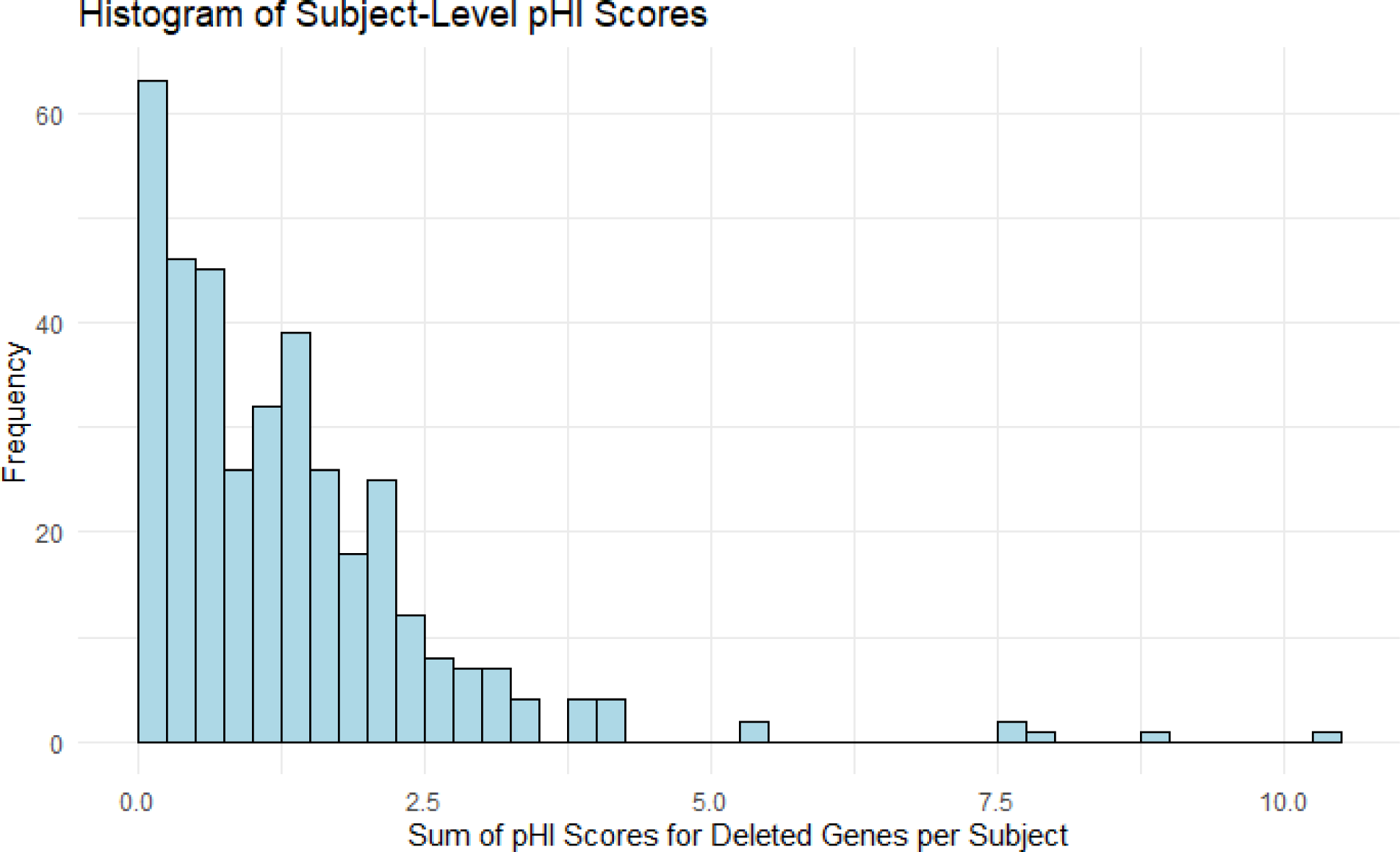
Distribution of sum of haploinsufficiency (pHI) scores for deleted genes per subject. 26 subjects had a sum of 0.

**Figure S15.**
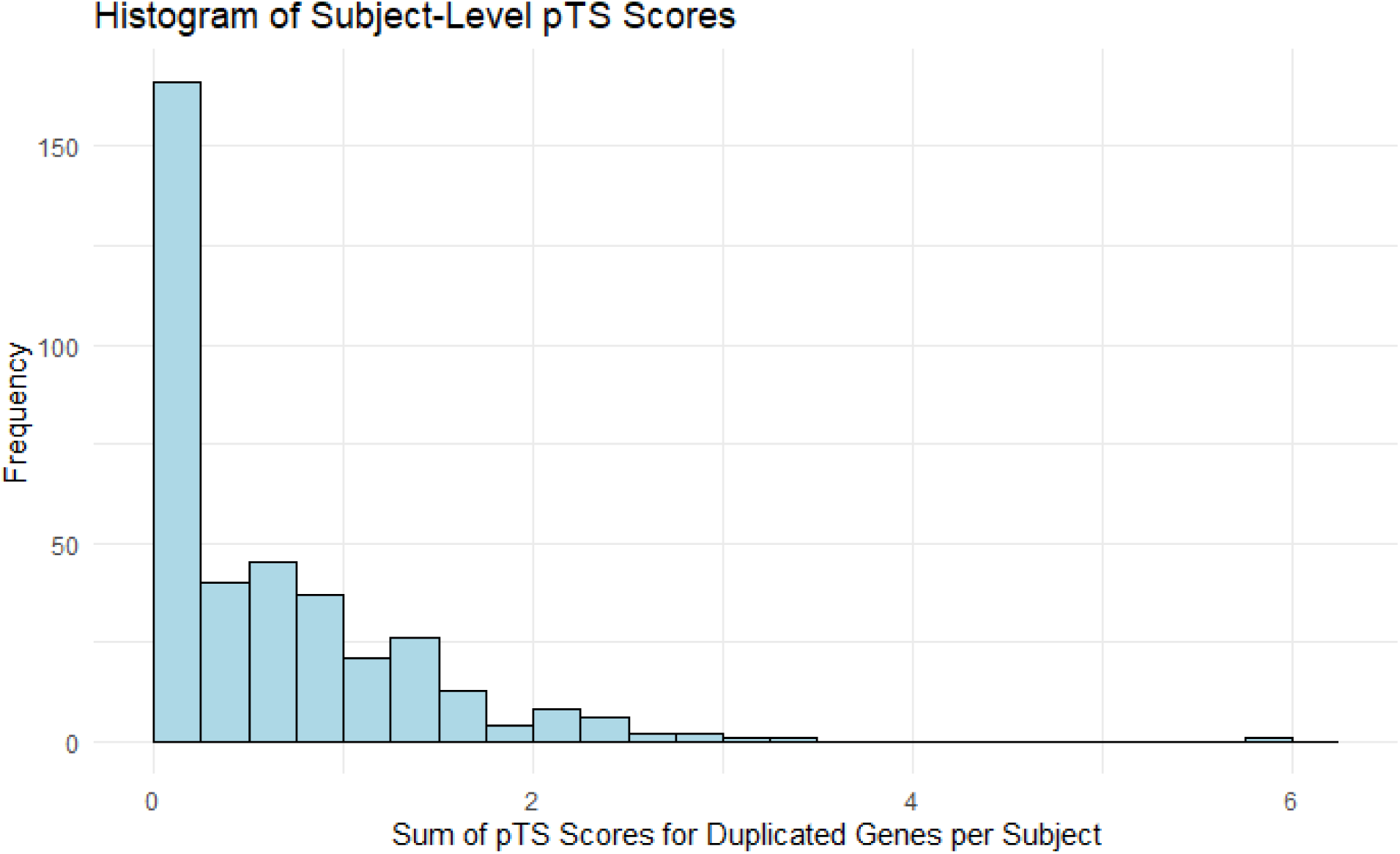
Distribution of sum of triplosensitivity (pTS) scores for duplicated genes per subject. 64 subjects had a sum of 0.

**Figure S16.**
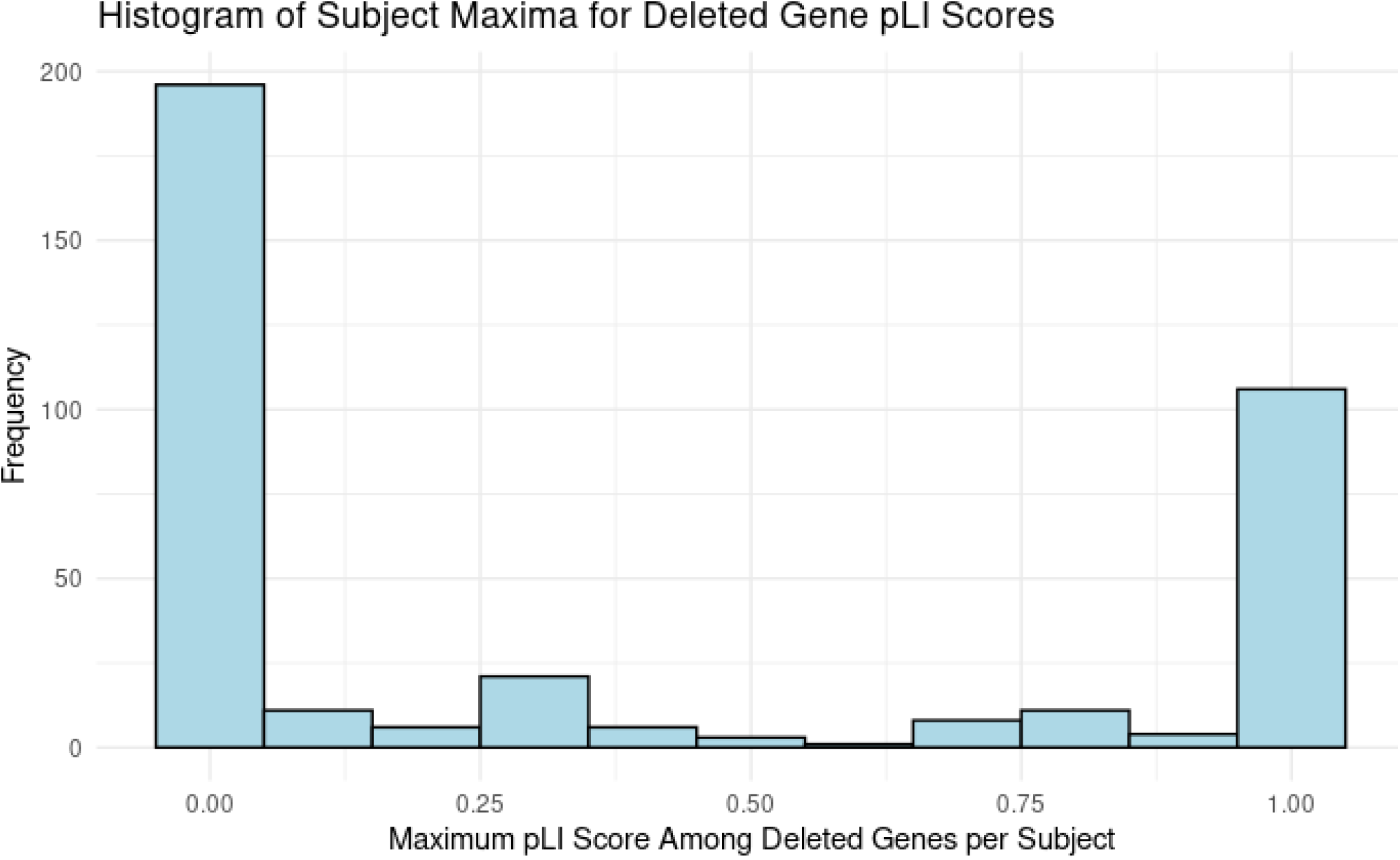
Distribution of maximum loss of function intolerance (pLI) score among deleted genes per subject.

**Figure S17.**
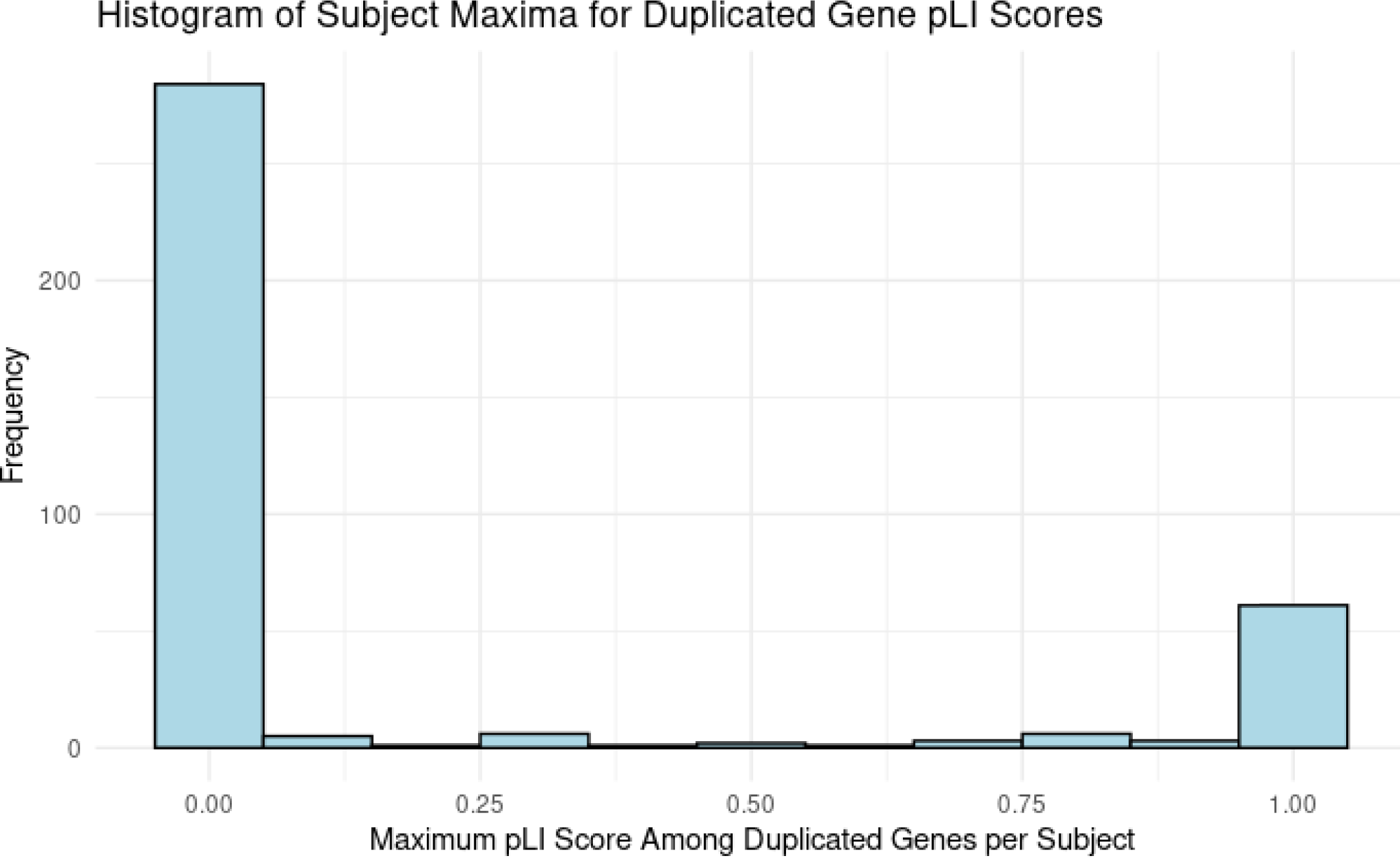
Distribution of maximum loss of function intolerance (pLI) score among duplicated genes per subject.

**Figure S18.**
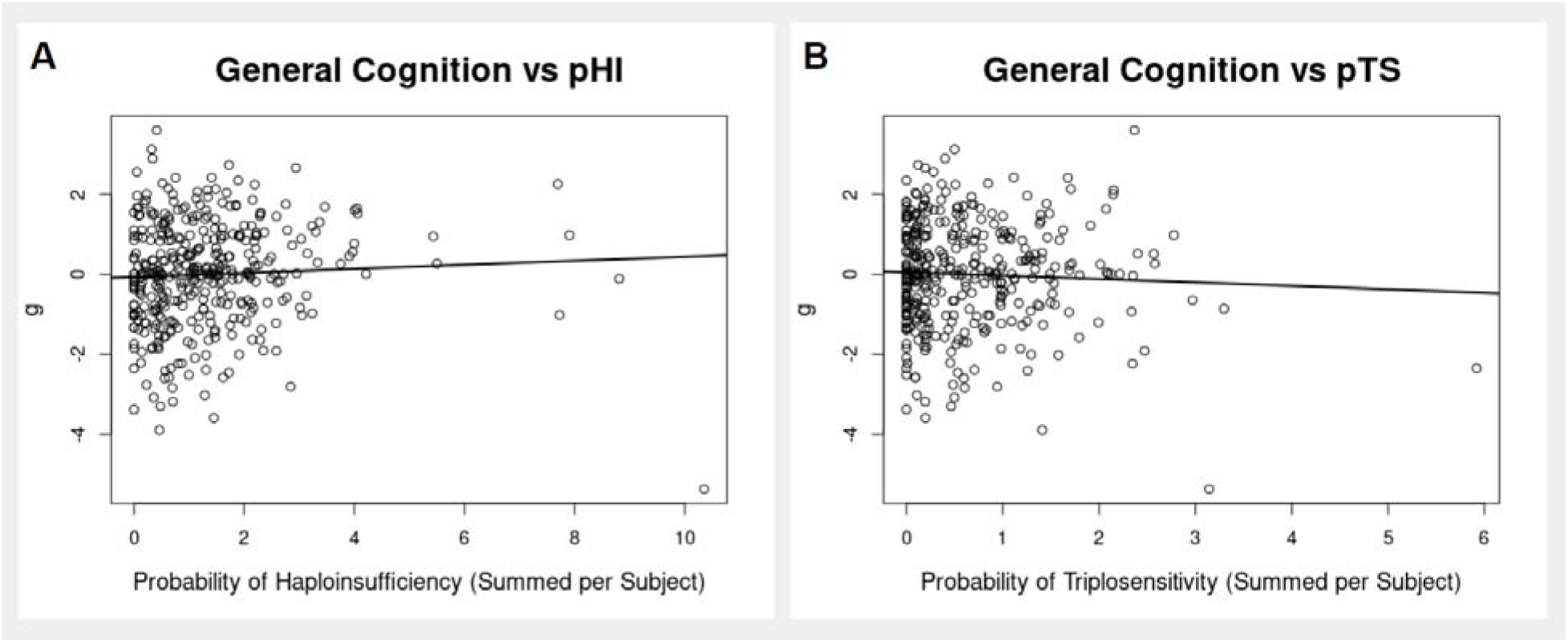
**A**. Scatter plot of general cognitive functioning vs. subject-level haplosensitivity (pHI) scores. Gene-level pHI scores were summed across all genes in deleted CNVs for each subject. Linear regression R2 = 0.0025, p = 0.34. **B**. Scatter plot of general cognitive functioning vs. subject-level triplosensitivity (pTS) scores. Gene-level pTS scores were summed across all genes in duplicated CNVs for each subject. Linear regression R2 = 0.0022, p = 0.37.

**Table S1.** Regression and correlation coefficients for each pair of cognitive tasks: L-values, adjusted R^2^-values and *p*-values.

|  |  |  |  |  |
| --- | --- | --- | --- | --- |
| Trails A | $\beta = -0.298$<br>Adj-R<sup>2</sup> = 0.090<br>$p = <0.001$ | x | x | x |
| Digit | $\beta = 0.319$<br>Adj-R<sup>2</sup> = 0.098<br>$p = <0.001$ | $\beta = -0.466$<br>Adj-R<sup>2</sup> = 0.202<br>$p = <0.001$ | x | x |
| VLT | $\beta = 0.325$<br>Adj-R<sup>2</sup> = 0.103<br>$p = <0.001$ | $\beta = -0.187$<br>Adj-R<sup>2</sup> = 0.034<br>$p = <0.001$ | $\beta = 0.337$<br>Adj-R<sup>2</sup> = 0.119<br>$p = <0.001$ | x |
| DANVA2 | $\beta = 0.203$<br>Adj-R<sup>2</sup> = 0.037<br>$p = <0.001$ | $\beta = -0.182$<br>Adj-R<sup>2</sup> = 0.030<br>$p = 0.002$ | $\beta = 0.212$<br>Adj-R<sup>2</sup> = 0.044<br>$p = <0.001$ | $\beta = 0.126$<br>Adj-R<sup>2</sup> = 0.013<br>$p = 0.022$ |
|  | Matrices | Trails A | Digit | VLT |

**Table S2.**
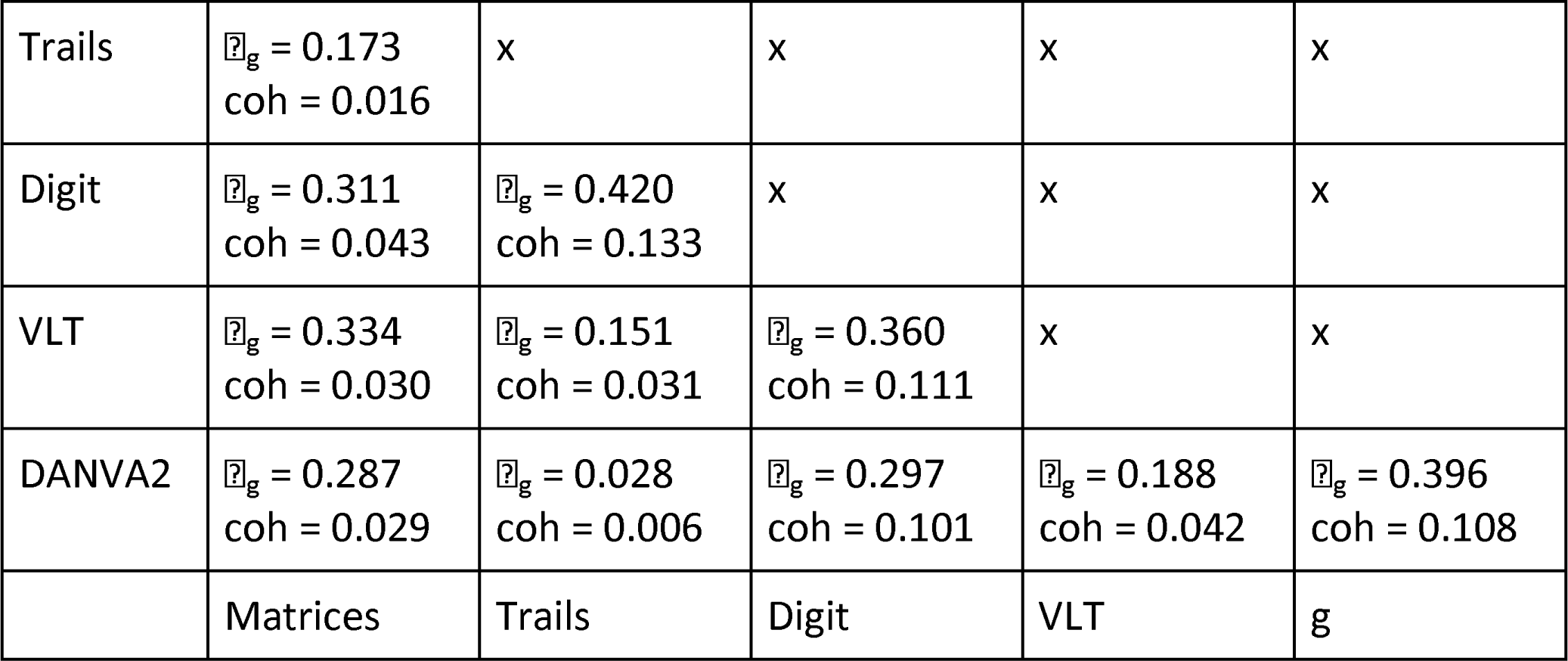
Genetic correlation (⍴_g_) and coheritability values (coh) between cognitive measures.

**Table S3.** Results of linear regressions between cognitive measures and genetic proximities of healthy subjects to the narrow and broad affected diagnostic categories. All p > 0.05.

| Cog. Measure | Matrices | Trails | Digit | VLT | DANVA2 | g |
| --- | --- | --- | --- | --- | --- | --- |
| Adj. R<sup>2</sup> - Prox. Narrow | -0.002 | 0.002 | -0.004 | -0.004 | -0.003 | -0.002 |
| *p-value* | 0.552 | 0.232 | 0.965 | 0.999 | 0.761 | 0.509 |
| Adj. R<sup>2</sup> - Prox. Broad | -0.003 | -0.001 | -0.005 | -0.003 | -0.001 | -0.004 |
| *p-value* | 0.586 | 0.391 | 0.875 | 0.560 | 0.380 | 0.841 |

**Table S4.** Adjusted R^2^ values for linear regressions of subject-level CNV risk scores against general cognition (g) and against general cognition compared to family average (compg). pHI sum = sum of probability of haploinsufficiency. pTS sum = sum of probability of triplosensitivity. pLI Del = maximum probability of loss of function intolerance among deleted genes. pLI Dup = maximum probability of loss of function intolerance among duplicated genes. Direction of correlation between g and pLI Del was positive (β = 0.3601).

| **CNV Score** | **pHI Sum** | **pTS Sum** | **pLI Del** | **pLI Dup** |
| --- | --- | --- | --- | --- |
| Adj R<sup>2</sup> - g | ≤0 | ≤0 | 0.012 | ≤0 |
| *p-value* | 0.336 | 0.372 | 0.021 | 0.535 |
| Adj R<sup>2</sup> - compg | ≤0 | ≤0 | ≤0 | ≤0 |
| *p-value* | 0.712 | 0.357 | 0.633 | 0.571 |

**Table S5.**
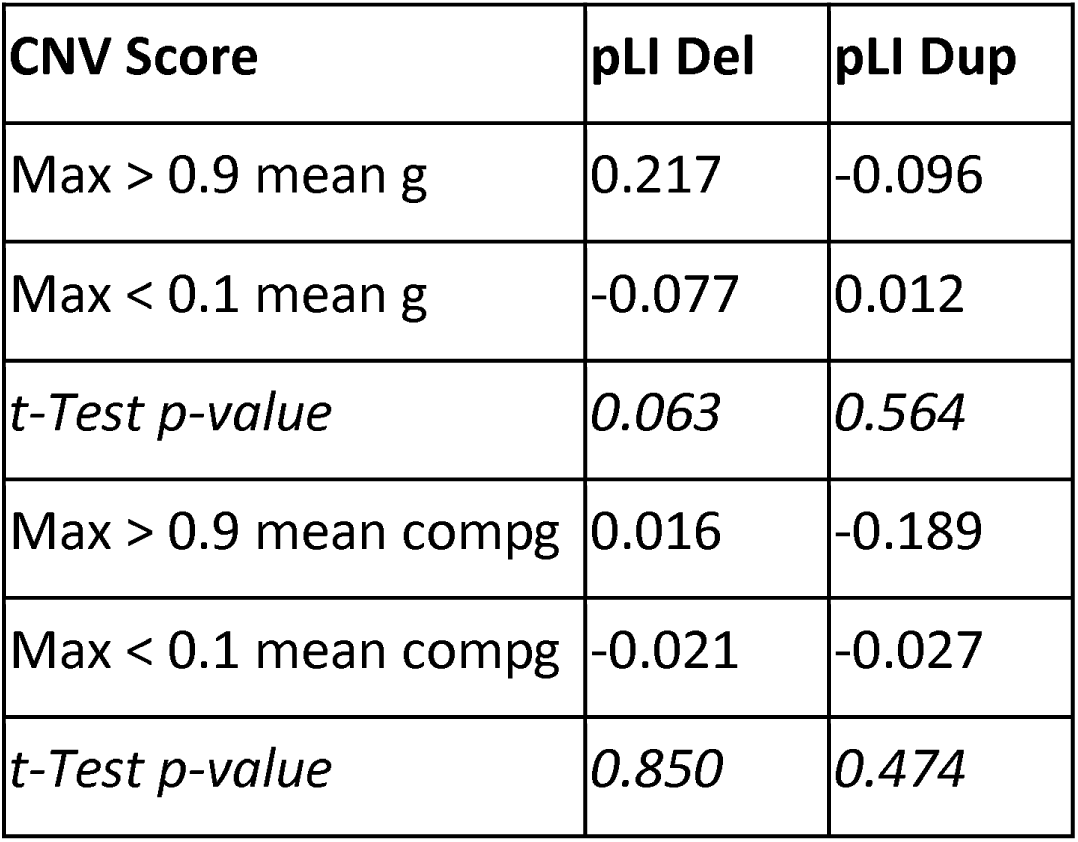
Means and Welch’s t-test p-values for general cognition (g), and general cognition compared to family average (compg), in subjects at extremes of max pLI score range (max pLI < 0.1 or max pLI > 0.9), for both deleted and duplicated genes. pLI Del = maximum probability of loss of function intolerance among deleted genes. pLI Dup = maximum probability of loss of function intolerance among duplicated genes.

